# Fourteen-days Evolution of COVID-19 Symptoms During the Third Wave in Non-vaccinated Subjects and Effects of Hesperidin Therapy: A randomized, double-blinded, placebo-controlled study

**DOI:** 10.1101/2021.10.04.21264483

**Authors:** Jocelyn Dupuis, Pierre Laurin, Jean-Claude Tardif, Leslie Hausermann, Camille Rosa, Marie-Claude Guertin, Karen Thibaudeau, Lyne Gagnon, Frank Cesari, Martin Robitaille, John E. Moran

**Affiliations:** Montreal Heart Institute Research Center, Montreal, Quebec, Canada; Department of Medicine of Université de Montréal, Montreal, Quebec, Canada; Ingenew Pharmaceuticals, Montreal, Quebec, Canada; Montreal Health Innovations Coordination Center, Montreal, Quebec, Canada

**Keywords:** flavonoid, COVID-19, viral infection, therapy, hesperidin, symptom

## Abstract

COVID-19 symptoms can cause substantial disability, yet no therapy can currently reduce their frequency or duration. We conducted a double-blind placebo-controlled trial of hesperidin 1000 mg once-daily for 14 days in 216 symptomatic non-vaccinated COVID-19 subjects. Thirteen symptoms were recorded after 3, 7, 10 and 14 days. The primary endpoint was the proportion of subjects with any of four cardinal (group A) symptoms: fever, cough, shortness of breath or anosmia. At baseline, symptoms in decreasing frequency were: cough (53.2%), weakness (44.9%), headache (42.6%), pain (35.2%), sore throat (28.7%), runny nose (26.9%), chills (22.7%), shortness of breath (22.2%), anosmia (18.5%), fever (16.2%), diarrhea (6.9%), nausea/vomiting (6.5%) and irritability/confusion (3.2%). Group A symptoms in the placebo vs hesperidin group was 88.8% vs 88.5% (day 1) and reduced to 58.5 vs 49.4 % at day 14 (OR 0.69, 95% CI 0.38–1.27, p = 0.23). At day 14, 15 subjects in the placebo group and 28 in the hesperidin group failed to report their symptoms. In an attrition bias analysis imputing “no symptoms” to missing values, the hesperidin group shows reduction of 14.5 % of group A symptoms from 50.9% to 36.4% (OR: 0.55, 0.32–0.96, p = 0.03). Anosmia, the most frequent persisting symptom (29.3%), was lowered by 7.3% at 25.3 % in the hesperidin group vs 32.6% in the placebo group (p = 0.29). Mean number of symptoms in placebo and hesperidin was 5.10 ± 2.26 vs 5.48 ± 2.35 (day 1) and 1.40 ± 1.65 vs 1.38 ± 1.76 (day 14) (p = 0.92). In conclusion, most non-vaccinated COVID-19 infected subjects remain symptomatic after 14 days with anosmia being the most frequently persisting symptom. Hesperidin 1g daily may help reduce group A symptoms. Earlier treatment of longer duration and/or higher dosage should be tested.

## Introduction

Since the end of December 2019, the COVID-19 pandemic led to important worldwide morbidity and mortality. Despite the success of vaccination, a substantial proportion of the world population is still awaiting immunization and therefore at risk of getting infected with the inherent risk of viral mutations that could lead to vaccine resistant strains of the virus. Most infected subjects report symptoms of varying severity that can become debilitating and persist for prolonged periods in a substantial proportion. Currently, no therapy that has been shown to reduce the burden and length of COVID-19 symptoms in non-hospitalized subjects.

COVID-19 is due to an infection by a novel beta-coronavirus, identified as 2019-nCoV [1], now known as Severe Acute Respiratory Syndrome – Coronavirus 2 (SARS-CoV-2), whose entry into cells has been shown to be dependant of the angiotensin converting enzyme 2 (ACE2) [2]. In a meta-analysis of 212 studies, Lie et al. [3] reported that the most common symptoms of COVID-19 were fever (78.8%), cough (53.9%) and malaise (37.9%). Other reported symptoms were fatigue (32.3%), expectoration (24.2%), myalgia (21.3%), shortness of breath (18.99%), chills (15.7%), diarrhea (9.5%), chest pain (9%), rhinorrhea (7.5%), vomiting (4.7%) and abdominal pain (4.5%). Furthermore, patients with a severe form of the disease were more subject to shortness of breath, abdominal pain, chills and dizziness than patients with a non-severe form. Studies also reported taste and smell dysfunction, such as anosmia, as a common symptom in people infected with COVID-19 [4, 5]. Some subjects at higher risk may show marked inflammation in response to the infection, referred to as the cytokine storm, leading to greater disease severity with acute respiratory distress and risk of hospital admission and death [6, 7]. The evolution of COVID-19 symptoms in non-hospitalized and non-vaccinated subjects during the third wave of the pandemic has not been reported. Compared to the first and second waves, large scale PCR testing became available during the third wave and was largely publicised and encouraged in all possibly infected subjects. The true proportion and evolution of symptoms may therefore differ from what was reported in previously more selected populations.

Hesperidin, a flavonoid naturally present in the peel of citrus fruits inhibits the 3-chymotrypsin like protease 3 (3CLpro) involved in SARS-CoV2 replication [8]. As well, hesperidin was reported to target the binding interface between the Spike protein of SARS-CoV-2 and the ACE2 receptor, potentially preventing the interaction of ACE 2 with the Spike regional binding domain (RBD) [9].

*In vivo* experimentation in rats infected with the H1N1 virus revealed that hesperidin effectively reduced lung impairment and suppressed pulmonary inflammation by reducing pro-inflammatory cytokines and recruitment of pro-inflammatory cells [10]. These anti-inflammatory and pulmonary protective effects were also reported in rats and mice with ventilator- and lipopolysaccharides-induced acute lung injury, respectively [11, 12]. Furthermore, evidence of cardioprotective and neuroprotective effects of flavanones, through their anti-oxidant and anti-inflammatory actions, were reviewed within the last decade and suggest the therapeutic potential of these compounds in conditions associated with inflammation and oxidative stress [13-15].

Considering its possible effects on SARS-CoV-2 entry into the cells and replication, as well as its anti-inflammatory action and its effectiveness in animal models of acute respiratory distress, hesperidin may be of interest in the treatment of COVID-19 related symptoms and complications. This study was designed to determine the effects of 14 days hesperidin treatment on the burden of COVID-19 symptoms in non-hospitalized subjects.

## METHODS

### Participants

This was a phase 2, randomized, double-blind, placebo-controlled study conducted at the Montreal Heart Institute (MHI) and comparing the effects of hesperidin (1000 mg once daily) and matching placebo (ratio 1:1) on COVID-19 symptoms during 14 days in participants infected with COVID-19 (detailed protocol is presented in S1 Protocol). Health Canada gave its authorization to conduct the study, which was also reviewed and approved by the Montreal Heart Institute Research and Ethics Committees (2021-2841). The study of hesperidin on COVID-19 symptoms (HESPERIDIN) was registered at clinicaltrials.gov (NCT04715932). A total of 216 subjects were recruited between February 18 and May 20, 2021 in the province of Quebec, Canada. All subjects were recruited by the Montreal Heart Institute research center and the study was coordinated by the Montreal Health Innovations Coordinating Center (MHICC).

Non-hospitalized male and female subjects of at least 18 years of age with a positive diagnosis of COVID-19 confirmed by polymerase chain reaction (PCR) testing within the last 48 hours and with at least one COVID-19 symptom were included. Female participants had to be without childbearing potential (postmenopausal for at least one year or surgically sterile) or with childbearing potential and practicing at least one method of contraception. Subjects were excluded if they were currently hospitalized or under immediate consideration for hospitalization, currently in shock or with hemodynamic instability, or undergoing chemotherapy for cancer. Other exclusion criteria for participants were: unable to take their oral temperature daily; having received a least one dose of the COVID-19 vaccine; pregnant (or considering becoming pregnant during the study) or breastfeeding women; taking anticoagulant or antiplatelet medications; bleeding disorders; and within 2 weeks of received or planned surgery. People with a regular consumption of natural products containing more than 150 mg of hesperidin or regular consumption of more than one glass of orange juice per day were also excluded, as were subjects with known allergy to any of the medicinal and non-medicinal ingredients of the study drug.

This was a no-contact study with the screening, randomization and follow-ups at day 3, 7, 10 and 14 done exclusively by phone. All randomized subjects signed an electronic informed consent form using the DocuSign online service. The study medication and material were delivered to the patients’ home and included an oral electronic thermometer (Physio logic^©^ Acuflex Pro) and a daily symptoms log. Allocation was performed through a randomization list generated by the MHICC (blocks sequence was fixed with block size of 4) and provided to the MHI pharmacists who dispensed the medication (hesperidin or placebo) according to the list after randomization of the participants by the study coordinators, keeping participants, investigators and staff blinded to drug assignment for the whole study duration. The symptoms log listed 13 COVID-19 symptoms including the temperature readings in degrees Celsius. Participants were asked to take two capsules (500 mg each) of study medication once daily at bedtime on an empty stomach. They were requested to record their symptoms and temperature daily in the symptoms log and return it to the study team at the end of their participation. At each follow-up call, the information recorded in the symptoms log was captured in an electronic case report form (InForm V 6.0, Oracle Health Sciences) by the study team. The trial ended according to protocol, namely after last patient last visit. The hesperidin capsules and matching placebo were kindly provided by Valeo Pharma (Kirkland, Quebec, Canada).

### Outcomes

The primary endpoint was the proportion of subjects with any of the following cardinal COVID-19 symptoms: fever, cough, shortness of breath or anosmia at day 3, 7, 10 and 14. These symptoms are referred to as group A symptoms in the province of Quebec, Canada. The secondary endpoints were: 1) The mean number of all COVID-19 symptoms (range 0-13) at day 3, 7, 10 and 14; 2) Duration of COVID-19 symptoms, defined as the number of days between first symptom and complete disappearance of any symptom; 3) For each 13 COVID-19 symptoms listed in the symptoms log (recent cough of aggravation of chronic cough, fever, chills, sore throat, runny nose, shortness of breath, nausea/vomiting, headache, general weakness, pain, irritability/confusion, diarrhea and anosmia defined as sudden loss of smell), the proportion of subjects with the symptom at day 3, 7, 10 and 14. Fever was defined as a temperature of > 38.0 °C by oral temperature using the supplied electronic thermometer.

The study also included two exploratory endpoints: 1) For each COVID-19 listed in the symptoms log, proportion of subjects with the symptom on a daily basis; and: 2) Composite of COVID-19 related hospitalization, mechanic ventilation or death in the 14 days following randomization.

The safety data were reviewed by a fully independent 3-member Data and Safety Monitoring Board (DSMB) after randomization of 50 subjects. Serious adverse events were reported to the DSMB on a weekly basis after their first meeting.

### Statistical analyses

Sample size was based on the proportion of subjects with any of the following group A COVID-19 symptoms: fever, cough, shortness of breath or anosmia at day 7. We assumed that 50% of placebo subjects would be symptomatic at this time point. These symptoms, referred to as group A symptoms in Quebec, are among the most frequent COVID-19 symptoms and are more objectively assessable. These symptoms were used as a diagnostic criterion in the epidemiological definition of COVID-19 prior to the widespread use and availability of PCR testing. Using a two-sided 0.05 significance level, considering achieving an 80% power to detect an absolute difference of 20% between both groups in the proportion of symptomatic participants, and factoring in a 15% drop out rate, we determined that 216 participants (108 per group) were required to complete the study.

Efficacy analyses were based on an intent-to-treat (ITT) principle. All participants who received the medication were included in the ITT population. The primary analysis compared the proportions of subjects between both treatment groups using a generalized linear mixed model (GLMM), more precisely, a repeated binary logistic regression model with terms for treatment group (placebo, hesperidin), time (3, 7, 10 and 14 days) and treatment group x time interaction. Contrasts under this model allowed for the comparisons of the proportions at each time point. Then, for secondary analyses, number of COVID-19 symptoms was compared between treatment groups using another GLMM, namely a repeated Poisson regression model with similar terms for group, time and interaction. Rate ratios are presented with 95% confidence intervals and p-values. Duration of COVID-19 symptoms were compared using a log rank test with Kaplan-Meier curves. Subjects who still had at least one symptom at their last assessment were censored at the day of this last assessment. The statistical approach used for primary endpoint was also used to compare individual COVID-19 symptoms over time. Composite of COVID-19-related hospitalization, mechanic ventilation or death were compared between both groups using a chi-square test. Statistical analyses were described in a statistical analysis plan that was approved prior to database lock and unblinding.

Safety of hesperidin was evaluated with descriptive statistics on adverse events and serious adverse events broken down by groups and presented on the safety population of all subjects who took at least one dose of the study medication.

To account for a possible attrition bias and evaluate its impact, two post-hoc sensitivity analyses on the primary endpoint were conducted. Both imputed data in subjects who stopped reporting symptoms prior to day 14. The first analysis used the last observation carried forward as a worst-case scenario to impute missing symptoms while the second imputed “no symptom” when symptoms were missing as a best-case scenario. All statistical tests were two-sided and conducted at the 0.05 significance level. Statistical analyses were done using SAS version 9.4.

## RESULTS

The study flow-chart is shown in figure 1. 217 subjects were enrolled and there was one screen-fail due to administration of COVID-19 vaccine prior to randomization. 216 subjects were randomized into the study with 109 assigned to placebo and 107 to hesperidin. All participants who received the placebo completed the study, but there was one lost to follow-up in the hesperidin group.

**Figure 1.**
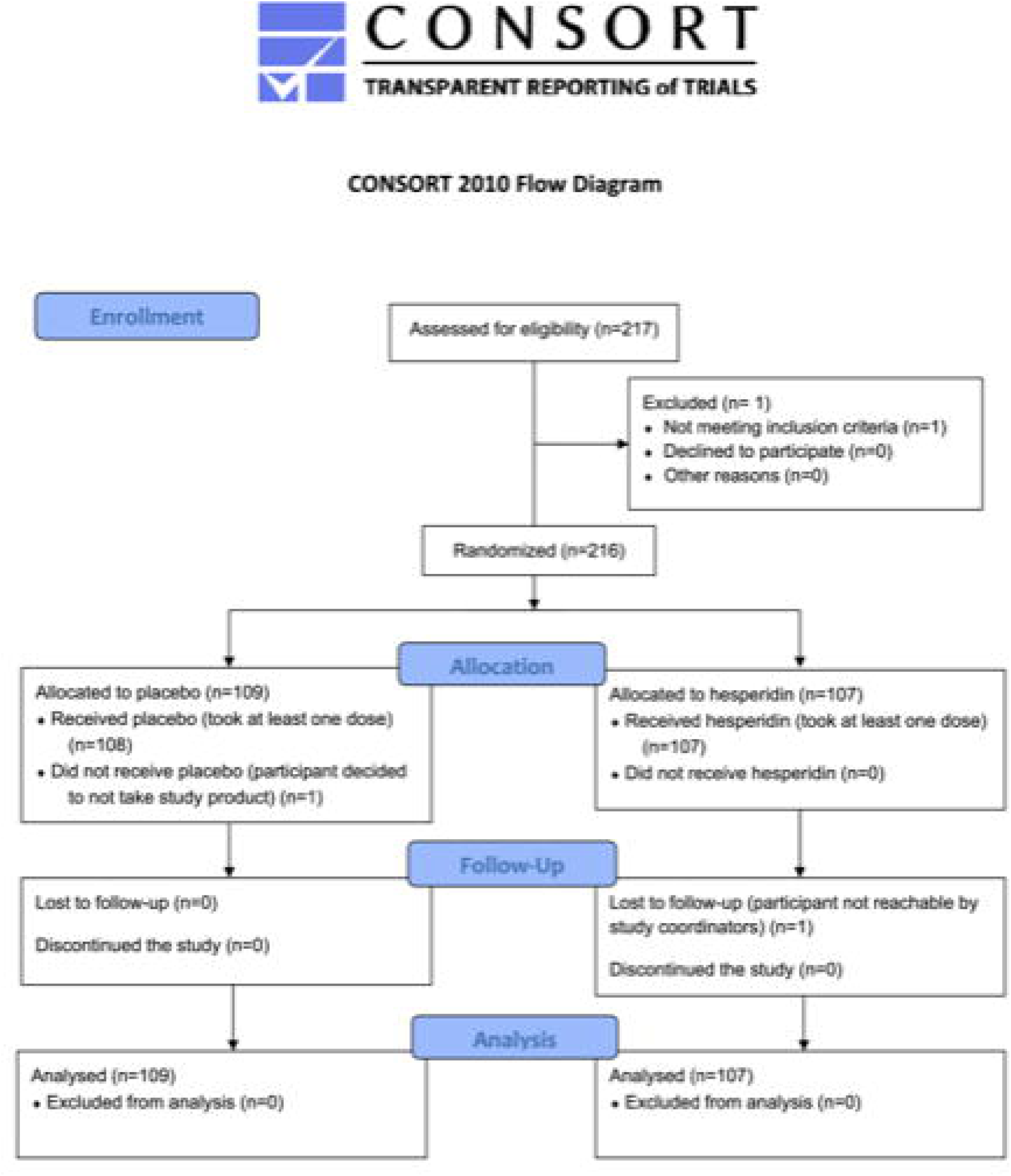
Study flow chart.

### Baseline characteristics (Table 1)

The demographics as well as the clinical profile at randomization are shown in table 1. For the whole study population, mean age was 40.98 ± 12.14 years with a proportion of males of 44.9%. The delay between the beginning of symptoms and randomization in the placebo group and the hesperidin group was similar at 3.78 ± 1.81 and 3.88 ± 1.89 days respectively. The mean delay between COVID-19 diagnosis and randomization was 1.10 ± 0.43 days in the placebo and 1.10 ± 0.39 days in the hesperidin group. The most common COVID-19 symptoms in decreasing frequency were: cough (53.2%), general weakness (44.9%), headache (42.6%), pain (35.2%), sore throat (28.7%), runny nose (26.9%), chills (22.7%), shortness of breath (22.2%), anosmia (18.5%), fever (16.2%), diarrhea (6.9%), nausea/vomiting (6.5%) and irritability/confusion (3.2%). This was a low-risk population evidenced by the low prevalence of diabetes, hypertension, heart diseases and respiratory diseases.

**Table 1.**
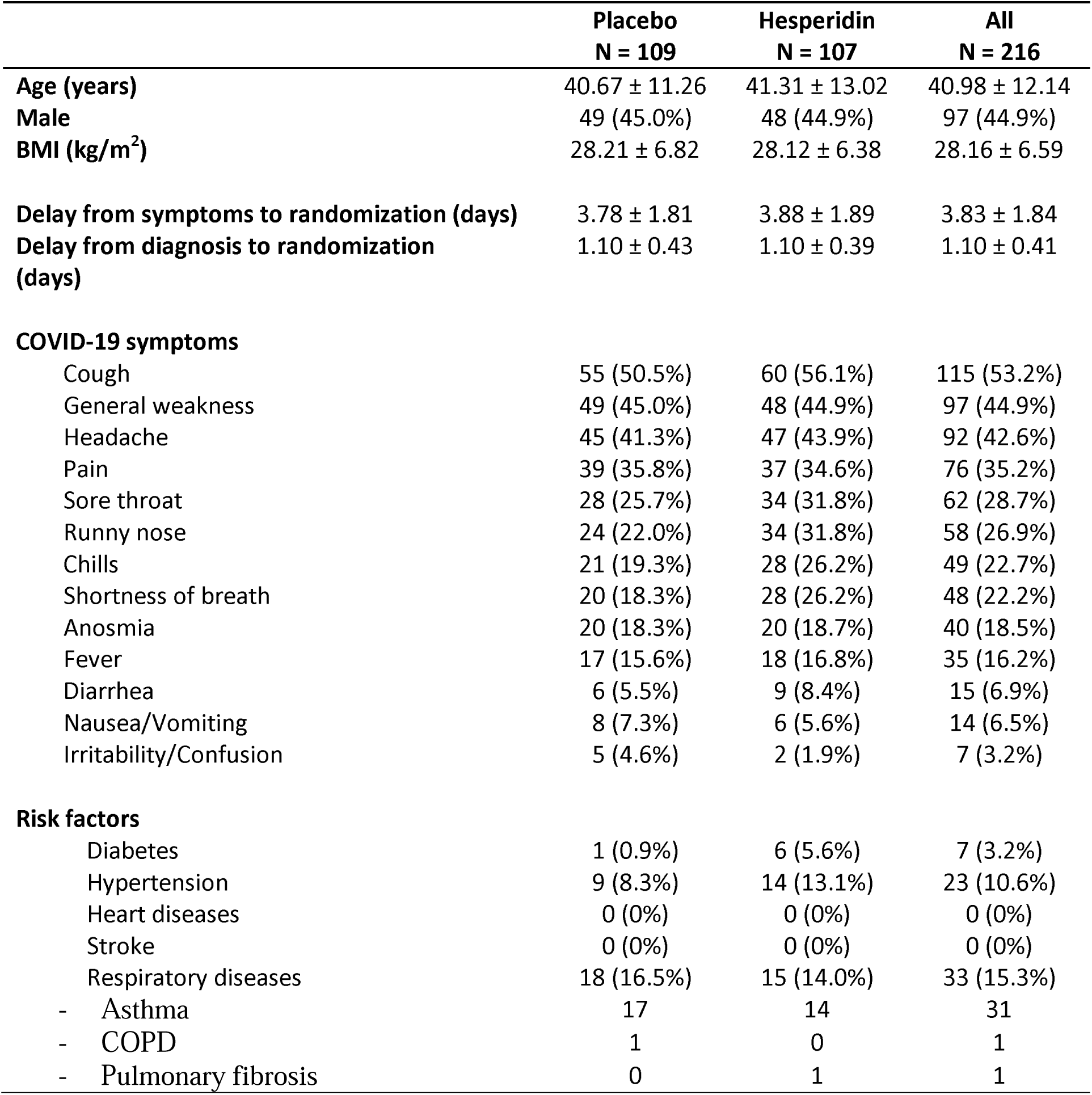
Patients’ baseline characteristics, intent-to-treat population.

|  | **Placebo**<br>**N = 109** | **Hesperidin**<br>**N = 107** | **All**<br>**N = 216** |
| --- | --- | --- | --- |
| **Age (years)** | 40.67 ± 11.26 | 41.31 ± 13.02 | 40.98 ± 12.14 |
| **Male** | 49 (45.0%) | 48 (44.9%) | 97 (44.9%) |
| **BMI (kg/m<sup>2</sup>)** | 28.21 ± 6.82 | 28.12 ± 6.38 | 28.16 ± 6.59 |
| **Delay from symptoms to randomization (days)** | 3.78 ± 1.81 | 3.88 ± 1.89 | 3.83 ± 1.84 |
| **Delay from diagnosis to randomization (days)** | 1.10 ± 0.43 | 1.10 ± 0.39 | 1.10 ± 0.41 |
| **COVID-19 symptoms** |  |  |  |
| Cough | 55 (50.5%) | 60 (56.1%) | 115 (53.2%) |
| General weakness | 49 (45.0%) | 48 (44.9%) | 97 (44.9%) |
| Headache | 45 (41.3%) | 47 (43.9%) | 92 (42.6%) |
| Pain | 39 (35.8%) | 37 (34.6%) | 76 (35.2%) |
| Sore throat | 28 (25.7%) | 34 (31.8%) | 62 (28.7%) |
| Runny nose | 24 (22.0%) | 34 (31.8%) | 58 (26.9%) |
| Chills | 21 (19.3%) | 28 (26.2%) | 49 (22.7%) |
| Shortness of breath | 20 (18.3%) | 28 (26.2%) | 48 (22.2%) |
| Anosmia | 20 (18.3%) | 20 (18.7%) | 40 (18.5%) |
| Fever | 17 (15.6%) | 18 (16.8%) | 35 (16.2%) |
| Diarrhea | 6 (5.5%) | 9 (8.4%) | 15 (6.9%) |
| Nausea/Vomiting | 8 (7.3%) | 6 (5.6%) | 14 (6.5%) |
| Irritability/Confusion | 5 (4.6%) | 2 (1.9%) | 7 (3.2%) |
| **Risk factors** |  |  |  |
| Diabetes | 1 (0.9%) | 6 (5.6%) | 7 (3.2%) |
| Hypertension | 9 (8.3%) | 14 (13.1%) | 23 (10.6%) |
| Heart diseases | 0 (0%) | 0 (0%) | 0 (0%) |
| Stroke | 0 (0%) | 0 (0%) | 0 (0%) |
| Respiratory diseases | 18 (16.5%) | 15 (14.0%) | 33 (15.3%) |
| - Asthma | 17 | 14 | 31 |
| - COPD | 1 | 0 | 1 |
| - Pulmonary fibrosis | 0 | 1 | 1 |

### Primary endpoint: proportion of participants with group A symptoms at day 3, 7, 10 and 14 (Table 2)

The proportion of subjects presenting with any of the four selected group A symptoms (fever, cough, shortness of breath and anosmia) in the hesperidin group compared to the placebo group were, respectively, 88.5% vs 88.8% (day 1), 91.2% vs 87.4% (day 3), 81.3% vs 75.2% (day 7), 64.4% vs 60.6% (day 10) and 49.4% vs 58.5% (day 14). At 14 days, there was a 9.1% absolute reduction in group A symptoms in the hesperidin group (OR: 0.69, p = 0.2328). There was progressive attrition in the number of participants that reported their symptoms between day 1 and day 14, with 15 missing in the placebo group and 28 in the hesperidin group. In the first post-hoc sensitivity analysis using the last observation carried forward imputation, there was also no statistically significant difference in the primary endpoint at each time point (S2 table). In this worst-case analysis, we still observed a reduction at day 14 in the hesperidin subjects from 59.3% to 52.3%, a 7.0 % difference (OR 0.75, p = 0.3098). In the second post-hoc sensitivity analysis imputing “no symptom” to any missing value (S2 table), the hesperidin group shows a statistically significant absolute reduction of 14.5 % of group A symptoms from 50.9% to 36.4% (OR: 0.55, p = 0.0343).

**Table 2.** Proportion of patients with group A COVID-19 symptoms, intent-to-treat population.

|  | Placebo | Hesperidin | All | OR (95% CI)<sup>a</sup> | P-value<sup>a</sup> |
| --- | --- | --- | --- | --- | --- |
| **Day 1** | N = 107 | N = 104 | N = 211 |  |  |
| **No** | 12 (11.2%) | 12 (11.5%) | 24 (11.4%) |  |  |
| **Yes<sup>b</sup>** | 95 (88.8%) | 92 (88.5%) | 187 (88.6%) | 0.97 (0.41; 2.28) | 0.9413 |
| **Day 3** | N = 103 | N = 102 | N = 205 |  |  |
| **No** | 13 (12.6%) | 9 (8.8%) | 22 (10.7%) |  |  |
| **Yes<sup>b</sup>** | 90 (87.4%) | 93 (91.2%) | 183 (89.3%) | 1.49 (0.60; 3.69) | 0.3849 |
| **Day 7** | N = 101 | N = 91 | N = 192 |  |  |
| **No** | 25 (24.8%) | 17 (18.7%) | 42 (21.9%) |  |  |
| **Yes<sup>b</sup>** | 76 (75.2%) | 74 (81.3%) | 150 (78.1%) | 1.43 (0.71; 2.88) | 0.3139 |
| **Day 10** | N = 99 | N = 90 | N = 189 |  |  |
| **No** | 39 (39.4%) | 32 (35.6%) | 71 (37.6%) |  |  |
| **Yes<sup>b</sup>** | 60 (60.6%) | 58 (64.4%) | 118 (62.4%) | 1.18 (0.65; 2.14) | 0.5886 |
| **Day 14** | N = 94 | N = 79 | N = 173 |  |  |
| **No** | 39 (41.5%) | 40 (50.6%) | 79 (45.7%) |  |  |
| **Yes<sup>b</sup>** | 55 (58.5%) | 39 (49.4%) | 94 (54.3%) | 0.69 (0.38; 1.27) | 0.2328 |

<sup>a</sup>Comparison between placebo group and hesperidin group. Significant when $p < 0.05$
<sup>b</sup>Subject has at least one of the group A COVID-19 symptoms: Fever, cough, shortness of breath or anosmia
N represents the number of subjects who completed the daily symptom diary.

### Effect of hesperidin treatment on the number of COVID-19 symptoms at day 3, 7, 10 and 14

Figure 2 presents the effect of hesperidin on the number of COVID-19 symptoms at day 1, 3, 7, 10 and 14. Hesperidin did not improve the mean number of COVID-19 symptoms for the whole treatment duration: day 1: 5.10 ± 2.26 vs 5.48 ± 2.35; day 3: 4.16 ± 2.39 vs 4.74 ± 2.52; day 7: 2.96 ± 2.46 vs 3.13 ± 2.49; day 10: 1.95 ± 2.12 vs 2.01 ± 2.19; day 14: 1.40 ± 1.65 vs 1.38 ± 1.76 in placebo vs hesperidin group respectively.

**Figure 2.**
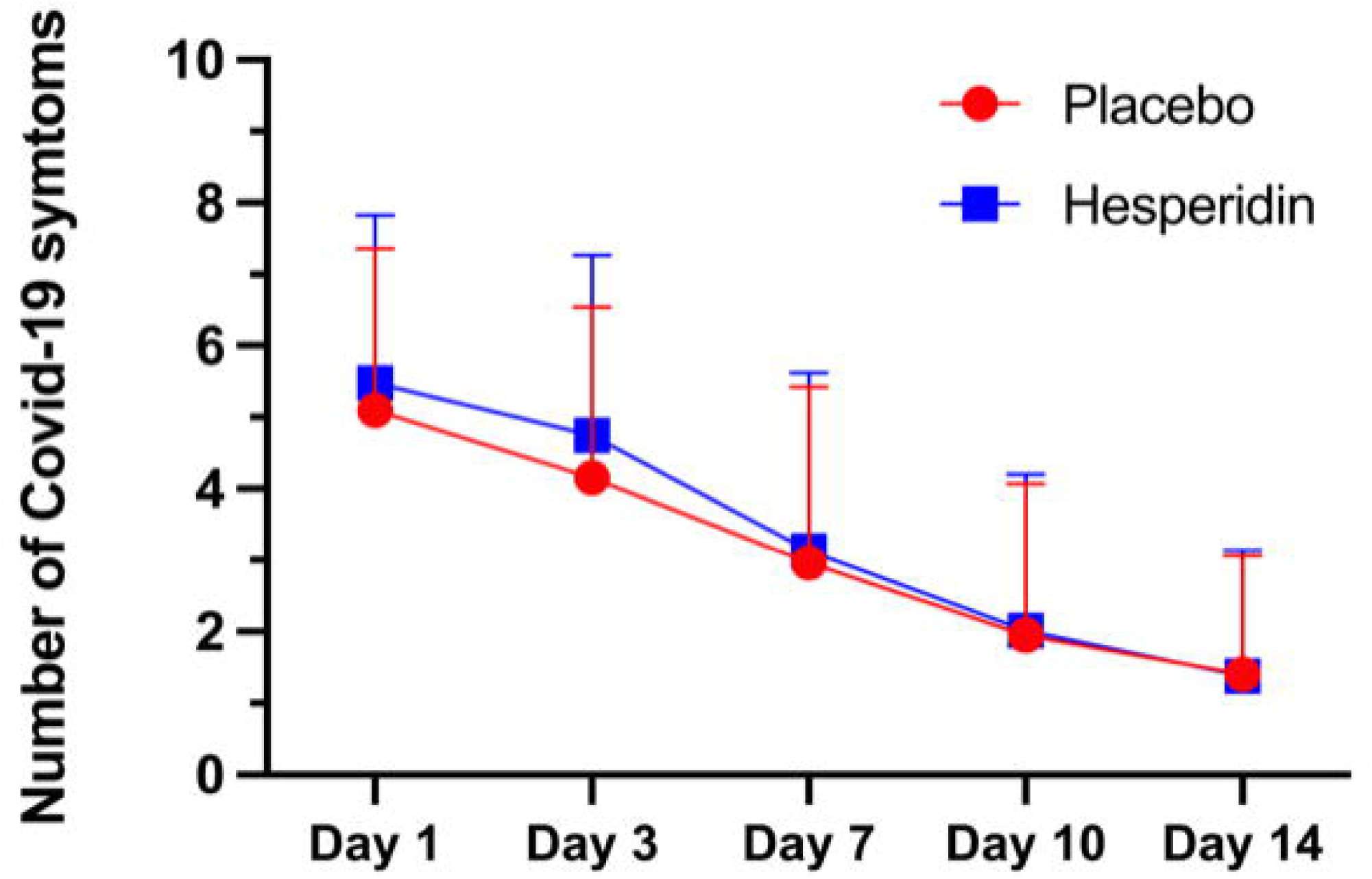
Mean number of COVID-19 symptoms at day 1, 3, 7, 10 and 14 in the placebo and the hesperidin groups. Values are mean ± SD.

### Effect of hesperidin treatment on the duration of COVID-19 symptoms

The Kaplan-Meier curve showing the proportion of symptom-free subjects over 14 days is shown in figure 3. Fourteen days post randomization, only 31.1% patients in the placebo group and 27.4% in the hesperidin group were symptom-free, indicating that the health of about 70% of our participants was still impacted by COVID-19 infection 14 days after study randomization and about 18 days after the beginning of symptoms. There was no difference in time to complete disappearance of symptoms between the two groups (p = 0.8834). In subjects with complete disappearance of symptoms, the duration of all COVID-19 symptoms in the placebo group vs the hesperidin group defined as the number of days between randomization and complete disappearance of any symptom was similar in both groups at 9.88 ± 2.71 days with placebo vs 10.34 ± 3.15 days with hesperidin.

**Figure 3:**
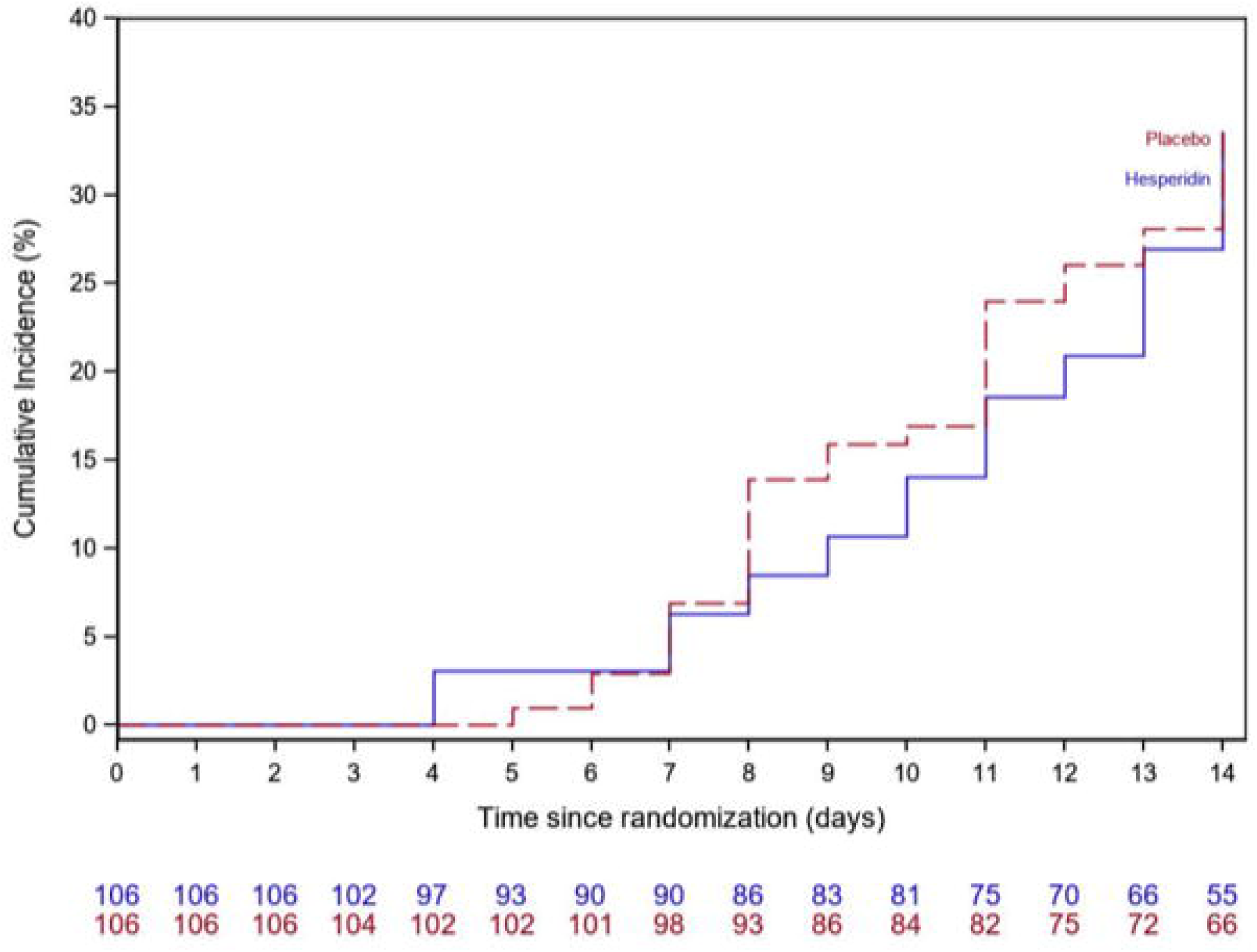
Kaplan-Meier curve. Proportion of symptom-free subjects over 14 days in the ITT population in the placebo and the hesperidin groups.

### Effect of hesperidin therapy on the proportion of subjects with each COVID-19 symptom and on the composite of hospital admission, mechanic ventilation and death

Figure 4 presents the proportion of subjects with each of the thirteen selected COVID-19 symptoms at day 1, 3, 7, 10 and 14. Detailed data and statistics for each symptom at each time point are presented in S3 tables. The four graphs in the top row of figure 4 represent the group A symptoms. The results showed that, except for fever which was absent at day 14 in the hesperidin group, each COVID-19 symptom was present at each time point in a certain proportion of patient that greatly varies depending on the symptom. For the whole duration of the study, the most prominent symptoms for their frequency and duration were cough and anosmia, two group A symptoms which affected 60.8% and 43.9% of participants at day 1 and persisted in 28.7% and 29.3% of them, respectively, at day 14. Some other symptoms such as runny nose, shortness of breath/difficulty breathing, headache and general weakness were still present in more than 10% of the whole population at the end of the study. All other symptoms were markedly reduced with time and only affected a small proportion of patient at day 14. For each time point, hesperidin had no statistically significant impact on the proportion of patients with each of these thirteen COVID-19 symptoms compared to the placebo group. Anosmia, the most frequently persisting symptom was the only symptom to increase during the time course of the study as 54.1% of the whole study group was affected at day 3. At day 14, persisting anosmia was reduced by 7.3% in the hesperidin group (25.3%) compared to the placebo group (32.6%, OR 0.70 [0.36 – 1.37], p = 0.2952).

**Figure 4.**
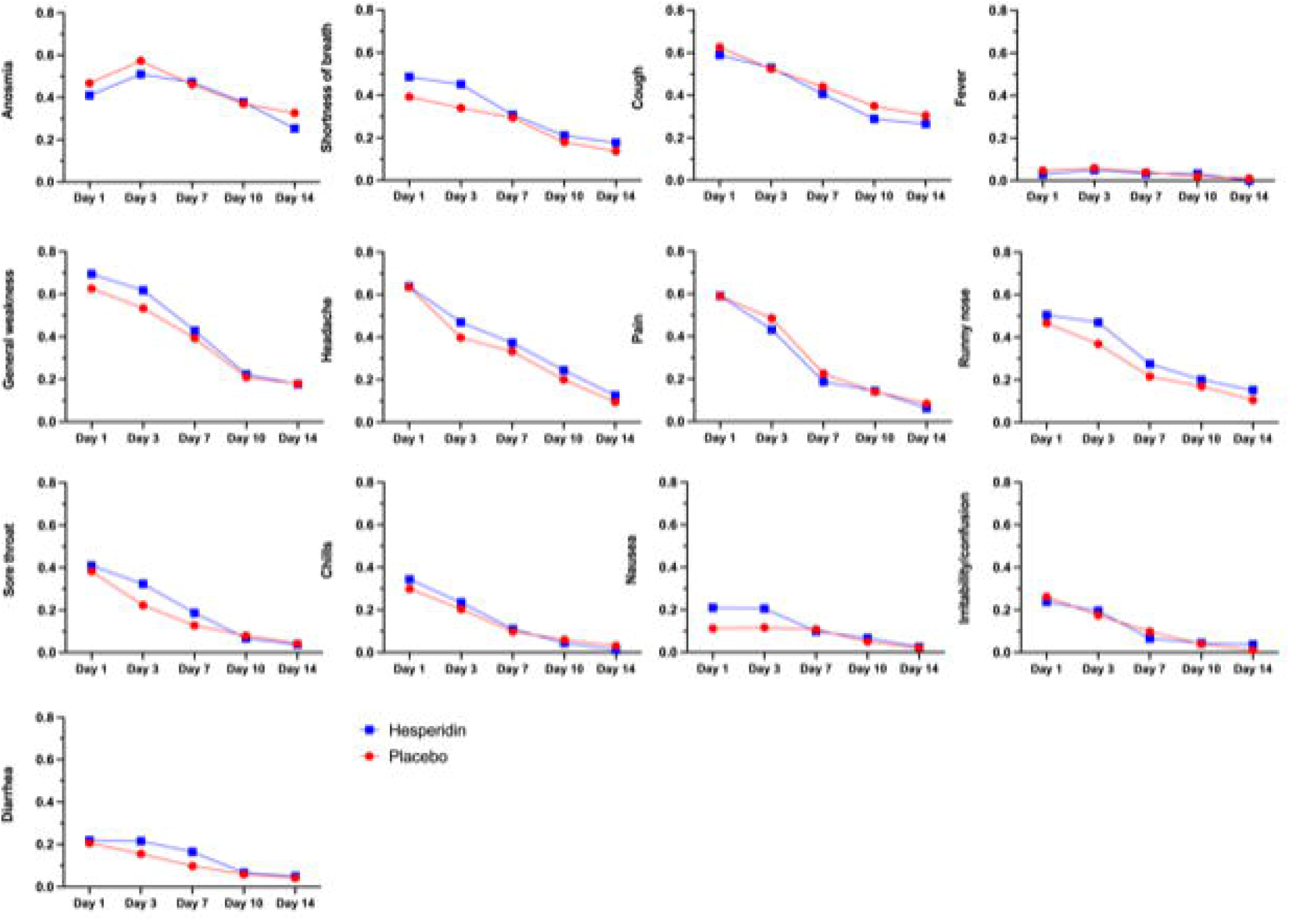
Proportion of subjects with each of thirteen COVID-19 symptoms at day 1, 3, 7, 10 and 14 in the placebo and the hesperidin groups.

The composite of COVID-19 related hospitalization, mechanic ventilation or death in the 14 days following randomization occurred in 1 subject in the placebo group and 3 subjects in the hesperidin group (p = 0.3669). One placebo subject was hospitalized for COVID-19 pneumonia that required intubation and ventilation. In the hesperidin group, 2 participants were hospitalized with pneumonia not requiring mechanical ventilation, and 1 subject was hospitalized for pneumonia and dehydration. There was no death.

### Safety profile of hesperidin

Treatment emergent adverse events (AE) and treatment emergent serious adverse events (TESAE) are presented in S4 table. There were 16 adverse events in the placebo group and 23 in the hesperidin group. Two subjects experienced at least one severe AE in the placebo group and 3 subjects in the hesperidin group. AE possibly related to study treatment occurred in 4 placebo participants and in 3 hesperidin participants. The majority of AE were related to COVID-19 infection. AE led to study drug withdrawal in 5 placebo subjects and 8 hesperidin subjects. There was 1 TESAE in the placebo group and 4 in the hesperidin group and none were related to study treatment.

## DISCUSSION

Commonly reported COVID-19 symptoms are cough, fever, malaise and anosmia [3, 4]. More severe cases present an exaggerated inflammatory response depicted as a cytokine storm that can lead to respiratory distress [6, 7]. Because SARS-CoV-2 is an evolutive virus with possible emergence of new variants that may not respond to current vaccines, it remains imperative to find treatments to reduce disease severity. Symptoms associated with COVID-19 in non-hospitalized subjects can be responsible for substantial disability, absenteeism, and loss of productivity [16]. During the first and second waves of the pandemic, availability of COVID-19 PCR diagnosis was limited and restricted to more symptomatic subjects, therefore introducing a selection bias in studies evaluating symptoms. During the recent third wave of the pandemic, PCR testing for COVID-19 became widely available and testing was strongly encouraged for all symptomatic subjects and contacts. There has been no prospective evaluation of COVID-19 symptoms in non-hospitalized and non-vaccinated subjects during the third wave. Here, we prospectively evaluated COVID-19 symptoms and the effects of 14-days hesperidin therapy, a flavonoid naturally present in citrus fruits, in 216 non-hospitalized and non-vaccinated symptomatic subjects who tested positive for COVID-19.

### Frequency and evolution of COVID-19 symptoms during the third wave

Subjects in this trial were randomized a mean of 3.83 ± 1.84 days after the beginning of symptoms and a mean of 1.10 ± 0.41 days after PCR diagnosis and followed for 14 days. Therefore, at the end of the study, the participants were at about 18 days since the beginning of symptoms. At randomization the most frequent symptoms, present in more than 1/3 of subjects, were cough, general weakness, headache and pain. In 20%-30% there was sore throat, runny nose, chills and shortness of breath.

Anosmia was present in 18.5% and fever in only 16.2%. Other symptoms, diarrhea, nausea/vomiting and irritability/confusion were present in only a minority of patients in a proportion of less than 7% each. With the notable exception of anosmia, all symptoms steadily decreased in frequency with time as the mean number of symptoms went from about 5.3 to 1.4 from day 1 to day 14. Still, most subjects, about 70%, remained symptomatic at day 14. The proportion of subjects with anosmia tripled from randomization to day 3 when it reached a proportion of 54.1%. At day 14, anosmia was the most frequent persisting symptom (29.3%). Considering the previous reports on the importance of persisting anosmia after COVID-19 and its impact on quality of life, our study confirms that anosmia occurs in about 50% of infected subjects and persists more than 14 days in about 30%. Clearly, because of its clinical importance, new sudden onset anosmia represents the best objective symptomatic target for COVID-19 therapeutic studies.

The incidence of fever found in this study is much lower that what has been previously reported early in the pandemic [17], but confirms later reports in non-hospitalized COVID-19 subjects that found comparable incidence [18]. Indeed, in 4066 outpatient adults with COVID-19 diagnosis and a mean age of 43, 10.3% reported fever [18]. This rate is similar to the 16.2% found at randomization in our study as self-reported by the participants. Our study further emphasizes the discrepancy in self-reported fever and mandatory measured temperature since we provided the subjects with a thermometer and required daily temperature measurements. Baseline temperature at randomization was inquired by phone, while day 1 temperature was measured with the provided electronic thermometer and entered in the symptoms log. Fever measured on day 1 in our study (defined as greater than 38.0 by oral thermometer) was present in only 3.9% of subjects, yet 32% reported chills on day 1 in the symptoms log. Our study therefore shows that objective fever is rare in most non-hospitalized COVID-19 subjects about 4 days after the beginning of symptoms.

### Effects of hesperidin therapy on COVID-19 symptoms

The primary endpoint of this trial was the proportion of subjects with any of 4 cardinal COVID-19 symptoms: fever, cough, shortness of breath and new onset anosmia. In the province of Quebec, Canada, they were referred to as group A symptoms, being more frequent and considered more specific for COVID-19 diagnosis. These symptoms were used for epidemiological diagnosis of COVID-19 contacts when large scale PCR testing was not available. Group A symptoms were present in 88.6% of patients on day 1 (88.8% placebo and 88.5% hesperidin) and persisted in 54.3% of patients on day 14. At day 14, hesperidin reduced group A symptoms by 8.9% from 58.5% in the placebo group to 49.4% without reaching statistical significance (OR 0.69, p = 0.23).

Despite repeated recalls by phone and emails, there was progressive attrition in the number of participants reporting symptoms, greater in the hesperidin group (28/107) than in the placebo group (15/109). To account and explore the extremes of a possible attrition bias, we performed a worst-case and best-case imputation analysis to missing values. In the worst-case analysis, we imputed the “last observation carried forward” approach and symptomatic subjects were therefore considered symptomatic for all subsequent missing days. In the best-case analysis, we imputed the absence of symptom to all missing values. In the worst-case analysis, we found no statistically significant difference in group A symptoms at all time points, but still observed a reduction at day 14 in the hesperidin group from 59.3% to 52.3%, a 7.0 % difference (OR 0.75, p = 0.3098). In the best-case analysis, the difference at day 14 became significant with a reduction of 14.5% from 50.9% in the placebo group to 36.4% in the hesperidin group (OR 0.55, p = 0.0343). Although speculative, the reason for the greater attrition rate in the reporting of symptoms in the hesperidin group may be due to symptomatic improvement and decreased willingness to cooperate for the participants that felt better. The attrition rate increased with study duration, a recognized factor of poorer compliance. Our study, powered to detect a 20% absolute difference in symptoms at Day 7, did not find statistically significant differences between treatments. A smaller absolute reduction, especially for anosmia, could however be highly clinically significant. Based on the attrition bias analysis and a best-case scenario where non-compliant subjects have no symptom, we cannot exclude that hesperidin could have beneficial effects and further studies are encouraged. Because of its clinical importance, persistence, and more subjective evaluation, new onset anosmia should be a primary therapeutic target in COVID-19 therapeutic studies.

The rationale and interest for using hesperidin in the treatment and even in the prevention of COVID-19 has been highlighted by others, both for its anti-oxidant and anti-inflammatory properties, and for its ability to block the entry and replication of SARS-CoV-2 [19, 20]. The current phase 2 study does not close the chapter on hesperidin therapy for COVID-19 with a signal of possible benefit on selected symptoms driven by a reduction of anosmia. Furthermore, since we did not grade the severity of each symptom in the design of this trial, we cannot exclude a potential benefit of treatment on this important component. Besides the attrition bias discussed above, there are several limitations that need to be considered in the planning of future phase 3 studies: delay of treatment, dosing, and duration of treatment and follow-up. The mean delay of 3.83 ± 1.84 days before enrollment into the trial may certainly mitigate the benefits of therapy as it has been largely reported that viral load peaks at symptoms onset and for the few following days, which is concordant with the infectiousness profile of COVID-19 [21]. The optimal therapeutic dosage of hesperidin has not previously been reported in human subjects. Participants were asked to take 2 capsules of 500 mg each once daily, the maximal allowable daily dose by the Non-Prescription and Natural Health Products Directorate (NNHPD) of Canada. Higher dosage more than once a day may be necessary to obtain optimal therapeutic effects. Finally, the duration of therapy and of follow-up may need to be longer to provide maximal benefit and better detect improvement of persisting symptoms, especially anosmia.

Our study showed good safety of hesperidin with no evidence for greater drug-related AE compared to placebo and no drug-related SAE. This concords with previous pre-clinical observations in Sprague Dawley rats, with low observed adverse effects at a dosage of 1000 mg/kg in a sub-chronic oral toxicity study [22]. As well, human studies showed a safe profile of hesperidin at a dosage ranging from 500 mg daily for 3 weeks [23] to 800 mg daily for up to 4 weeks [24] in both men and women. Although we excluded pregnant women from the current study, the use of veinotonics containing hesperidin to treat hemorrhoids and varices in pregnant women appears safe with no increase in reported adverse outcomes [25]. Finally, the US Food and Drug Administration issued a Generally Recognized as Safe Notice (GRAS No.796) in 2018 [26] for the use of orange extract with 85% hesperidin content as well as GRAS No. 901 [27] for glucosyl hesperidin to be used as additives in food and beverages. Collectively, these data support the use of higher dosage of hesperidin in future trials.

## Conclusion

During the third wave of the COVID-19 pandemic, only 30% of initially symptomatic non-hospitalized and non-vaccinated subjects were asymptomatic about 18 days after symptom onset. Anosmia affected 50% of subjects and was the most frequently persisting symptom in 30%. Hesperidin therapy is safe and may help reduce a composite of selected COVID-19 symptoms including fever, cough, shortness of breath and anosmia. Further trials with this agent are encouraged.

## Supporting information

S1 Protocol

S5 CONSORT 2010 Checklist

## Data Availability

All data are fully available without restriction

## Acknowledgements

The authors would like to thank: The Montreal Heart Institute Foundation for providing funding for this study; Sarah Samson RN, Denis Fortin RN, Cyntia Pennestri RN, Colette Morin RN, Natasha Moisan RN, Patricia-Iva Blaise RN, Nancy Fernandez RN, Jessica Drouin RN, Amélie Allard LPN, Ramona Badaluta RN, Myriam Duhamel RN, Marie-Gabrielle Lessard RN and Alexandre Bergeron from the Montreal Heart Institute clinical study team for their hard work in recruiting and following-up participants during the whole study; pharmacists Marie Robitaille, Lucie Verret, Sylvia Audet, Arnaud Canneva and Jeannie Meideros Charbonneau and pharmacy technicians from the Montreal Heart Institute pharmacy study team; the biostatistician and project management teams at the Montreal Health Innovations Coordinating Center for their expertise in development and project management, as well as statistical analyses; Chantal Lacoste for her help in setting up the telephone system for remote monitoring of participants; and the telephone operators who initiated first contact with the study participants; Alberto Mazza and Caroline Morneau for multiple insightful discussions on the scientific rational supporting the hesperidin clinical study; the Montreal Heart Institute Foundation for the financial support through donations by the Ariane Riou and Réal Plourde Foundation, the Sandra and Alain Bouchard Foundation, the Lise and Richard Fortin Foundation and Jacques D’Amours; Valeo Pharma Inc. for providing the study drug and placebo free of charge.

## Supporting information caption

**S1 Protocol. Version 2.0 of the Hesperidin study protocol approved by the Montreal Heart Institute Research and Ethics Committees**

**S2 table. Proportion of patients with group A COVID-19 symptoms – Attrition bias analysis**

**S3 table. Summary of and statistical analysis for the presence of each 13 COVID-19 symptoms**

**S4 table. Overall summary of treatment emergent adverse events (TEAE) and serious adverse events (TESAE)**

**S5 CONSORT 2010 checklist**.

