## Supplementary material for "Fourteen-days Evolution of COVID-19 Symptoms During the Third Wave in Non-vaccinated Subjects and Effects of Hesperidin Therapy: A randomized, double-blinded, placebo-controlled study": S1 Protocol

### CLINICAL RESEARCH PROTOCOL

#### Protocol # MHICC-2020-003

Final Protocol V2.0

February 17th, 2021

**SPONSOR**

MONTREAL HEART INSTITUTE

CONFIDENTIALITY

The Montreal Health Innovations Coordinating Center (MHICC), a division of the Montreal Heart Institute (MHI), considers the information contained in this document to be proprietary and/or confidential. It is for the use of the investigational team, and the corresponding applications to institutional review boards (IRBs) and independent ethics committees (IECs) in support of a clinical trial. Accordingly, please do not copy, use, distribute, or disclose to others without the written prior approval of MHICC or their designated representatives.

#### Signatures for:

### Protocol # MHICC-2020-003

**A phase 2, randomized, double-blind, placebo-controlled study of hesperidin therapy on COVID-19 symptoms:**
**The Hesperidin Coronavirus study (Hesperidin)**

| | | |
|---|---|---|
| Andreas Orfanos, MBBCh, FFPM, MBA<br>Director, MHICC | (Signature) | (Date) |
| Marie-Claude Guertin, Ph.D.<br>Head, Biostatistics Department, MHICC | (Signature) | (Date) |
| Jocelyn Dupuis, MD, Ph.D<br>Principal Investigator | (Signature) | (Date) |

8.3.3 Interim analysis ... 24
**9 STUDY COORDINATION ... 24**
**REFERENCES ... 25**
**LIST OF APPENDICES ... 27**
**Appendix A Detailed Study Safety Parameters ... 28**
**Appendix B Administrative Procedures for the Reporting of Adverse Events ... 29**
**1 ADVERSE EVENTS DURING THE TRIAL ... 29**
1.1 Pre-existing condition ... 29
1.2 Adverse Event (AE) ... 29
1.3 Related Adverse Event ... 29
1.4 Serious Adverse Event (SAE) ... 29
1.5 Life-Threatening Adverse Event ... 30
1.6 Clinical Laboratory Adverse Event ... 30
1.7 Treatment-Emergent Signs and Symptoms (TESS) ... 30
1.8 Post-treatment Adverse Event ... 30
**2 HANDLING OF ADVERSE EVENTS ... 31**
2.1 Treatment-Emergent Signs and Symptoms ... 31
2.2 Serious Adverse Events ... 31
2.3 Intensity ... 31
2.4 Relationship to Study Drug – Physician's Assessment ... 31
2.5 Clinical Outcome ... 31
**3 CAPTURING AVDERSE EVENTS ... 32**
3.1 Pre-Existing Condition ... 32
3.2 Clinical Laboratory Adverse Event ... 32
3.3 Hospitalization or Surgery/Procedure ... 32
3.4 Death ... 32
**4 REPORTING TO THE SPONSOR ... 32**
4.1 Immediately Reportable Adverse Events ... 32
4.2 Other Adverse Events ... 32
4.3 Follow-Up Period ... 33
**Appendix C Other Administrative and Regulatory Procedures ... 34**
**Appendix D Declaration of Helsinki ... 38**
**Appendix E ... 45**

#### INVESTIGATOR'S SIGNATURE

I have read the protocol and agree to conduct this trial in accordance with all stipulations of the protocol, with applicable laws and regulations and in accordance with the ethical principles outlined in the Declaration of Helsinki.

By signing below, I hereby declare that I am not debarred, disqualified, or otherwise restricted by any agency from conducting any research studies.

| Principal Investigator | Signature | Date |
| --- | --- | --- |

Country:

#### SYNOPSIS

**TITLE: A phase 2, randomized, double-blind, placebo-controlled study of hesperidin therapy on COVID-19 symptoms:**
**The Hesperidin Coronavirus study (Hesperidin)**

**INDICATION**: Reduction of symptoms in patients with COVID-19 infection.

**OBJECTIVES**:

- The **primary objective** of the study is to determine the effect of hesperidin therapy on the presence of selected COVID-19 symptoms (fever, cough, shortness of breath or anosmia) at 3, 7, 10 and 14 days following randomization.

- The **secondary objectives** are:
  1- To determine the effect of hesperidin therapy on the mean number of COVID-19 symptoms at 3, 7, 10 and 14 days following randomization.
  2- To determine the effect of hesperidin therapy on the duration of COVID-19 symptoms.
  3- To determine the effect of hesperidin therapy on the presence of each individual COVID-19 symptoms at 3, 7, 10 and 14 days following randomization.

- The **exploratory objectives** are:
  1- To evaluate the effect of hesperidin therapy on the presence of each individual COVID-19 symptoms on a daily basis based on a patient symptoms' diary.
  2- To evaluate the effect of hesperidin therapy on COVID-19-related hospitalization, mechanical ventilation and death in the 14 days following randomization.

**PATIENT POPULATION**: Males and females, at least 18 years of age, who have been diagnosed with a COVID-19 infection and have one or more symptoms.

Inclusion Criteria:

- Covid-19 positive by polymerase chain reaction (PCR) testing;

- Participant must be able to evaluate their symptoms and report them in the symptoms' diary;

- Patients must be able to take their oral temperature daily with an electronic thermometer provided to them with study materials.

- Males and females, at least 18 years of age, capable and willing to provide informed consent;

- Female patient is either not of childbearing potential, defined as postmenopausal for at least 1 year or surgically sterile, or is of childbearing potential and practicing at least one method of contraception and preferably two complementary forms of contraception including a barrier method (e.g. male or female condoms, spermicides, sponges, foams, jellies, diaphragm, intrauterine device (IUD)) throughout the study;

- Patient must have received a diagnosis of COVID-19 infection within the last 48 hours and have one or more symptoms;

- Outpatient setting (not currently hospitalized or under immediate consideration for hospitalization);

- Patient must be able and willing to comply with the requirements of this study protocol.

Exclusion Criteria:

- Patient currently hospitalized or under immediate consideration for hospitalization;

- Patient currently in shock or with hemodynamic instability;

- Patient undergoing chemotherapy for cancer;

- Patient is unable to take oral temperature using an electronic thermometer;

- Patient who received at least one dose of the COVID-19 vaccine;

- Female patient who is pregnant or breast-feeding or is considering becoming pregnant during the study;

- People taking anticoagulant/antiplatelet medications, those with bleeding disorders, and people two weeks before or after surgery;

- Patient is considered by the investigator, for any reason, to be an unsuitable candidate for the study;

- Regular consumption of natural products containing more than 150 mg of hesperidin or regular consumption of more than 1 glass of orange juice per day;

- Known allergy to any of the medicinal and non-medicinal ingredient: hesperidin, microcrystalline cellulose, magnesium stearate.

**STUDY DESIGN**: This will be a randomized, double-blind, placebo-controlled study. The study will include subjects from Quebec diagnosed with COVID-19 infections. Following informed consent, 216 subjects meeting all inclusion and no exclusion criteria will be randomized to receive either hesperidin 1000 mg once daily (q.d.)) or placebo (1:1 allocation ratio) for 14 days. Investigational drug will be delivered to the patients' homes with an electronic oral thermometer and a symptoms diary. Follow-up phone assessments will occur after 3, 7, 10, and 14 days following randomization for evaluation of COVID-19 symptoms. Electronic Case Report Form (eCRF) will be completed by the research personnel over the phone with the patients. The symptoms diary will be mailed back to the coordinating center at the end of the study in a pre-addressed envelope.

**OUTCOMES:**

- The **primary endpoint**:
  Proportion of subjects with any of the following COVID-19 symptoms: fever, cough, shortness of breath or anosmia, at day 3, 7, 10 and 14.

- The **secondary endpoints**:
  1- Mean number of COVID-19 symptoms (range 0-13) at day 3, 7, 10 and 14.
  2- Duration of COVID-19 symptoms, defined as the number of days between first symptoms and complete disappearance of any symptom.
  3- For each COVID-19 symptom listed in the appendix, proportion of subjects with the symptom at day 3, 7, 10 and 14.

- The **exploratory endpoints**:
  1- For each COVID-19 symptom listed in the appendix, proportion of subjects with the symptom on a daily basis based on patient-diary.
  2- Composite of COVID-19-related hospitalization, mechanic ventilation or death in the 14 days following randomization.

**STATISTICAL RATIONALE AND ANALYSIS:**

Efficacy analysis will be based on the intent-to-treat principle. The primary analysis will compare the proportions of subjects with fever, cough, shortness of breath or anosmia between the two treatment groups using a generalized linear mixed model with terms for treatment group (placebo, hesperidin), time (3, 7, 10 and 14 days) and treatment group x time interaction. The primary analysis will be the comparison between treatment groups at day 7, although treatment groups will also be compared at the other time points. Mean number of COVID-19 symptoms will be compared between treatment groups using a Student t-test or a Wilcoxon rank-sum test according to distribution. If more appropriate, a Poisson regression model will be used. Duration of COVID-19 symptoms will be compared between treatment groups using a log rank test and Kaplan-Meier curves will be presented. Subjects who will still have COVID-19 symptoms at the end of the study will be censored at the day of their last phone contact or videovisit. The analysis of individual COVID-19 symptoms over time will be similar to the analysis of the primary endpoint and will use generalized linear mixed models. Composite of COVID-19-related hospitalization, mechanic ventilation or death will be compared between treatment groups using a chi-square test. All statistical tests will be two-sided and conducted at the 0.05 significance level. Statistical analysis will be done using SAS version 9.4 or higher.

Using a two-sided 0.05 significance level, sample sizes of 91 in both groups (182 total) will achieve 80% power to detect an absolute between groups difference of 20% in the proportion of patients with COVID-19 symptoms (fever, cough, shortness of breath or anosmia) at day 7, which will be the time point of primary focus. This is assuming that the proportions of patients with the above COVID-19 symptoms at day 7 will be 50% in the placebo group and 30% in the active treatment group. Factoring in a 15% drop out rate, a total of 216 patients will be randomized in the study (108 per group).

A fully independent 3-member Data and Safety Monitoring Board (DSMB) will be established and will review unblinded safety data as detailed in the DSMB charter after randomization of 50 subjects and after randomization of 100 subjects. If necessary, an additional meeting will take place after randomization of 150 subjects. The DSMB charter will pre-specify the rules for early study termination, approved by all board members.

**ANTICIPATED TOTAL NUMBER OF RANDOMIZED PATIENTS**: 216 patients in total will be randomized in this study.

**PHASE**: 2

**STUDY LOCATION**: Canada

#### GLOSSARY

| | | |
|---|---|---|
| ACE2 | - | Angiotensin Converting Enzyme 2 |
| AE | - | Adverse event |
| eCRF | - | electronic Case Report Form |
| DSMB | - | Data and Safety Monitoring Board |
| EC | - | Ethics Committee |
| e-Consent | - | electronic Consent |
| ICF | | Informed Consent Form |
| ICH | - | International Council for Harmonisation |
| ITT | - | Intent-to-treat |
| IUD | - | Intrauterine device |
| IRB | - | Institutional Review Board |
| LPS | - | Lipopolysaccharides |
| MHI | - | Montreal Heart Institute |
| MHICC | - | Montreal Health Innovations Coordinating Center |
| PCR | - | Polymerase Chain Reaction |
| qPCR | - | quantitative Polymerase Chain Reaction |
| PO (po) | - | Per os |
| PRN | - | pro re nata (as necessary) |
| SAE | - | Serious adverse event |
| SOPs | - | Standard operating procedures |
| SUSAR | - | Suspected Unexpected Serious Adverse Reaction |
| TESS | - | Treatment-Emergent Signs and Symptoms |
| WMA | - | World Medical Association |

**TIME PERIOD AND NUMBER OF PATIENTS:**

A. Anticipated Starting Date of Study: 3Q20
B. Anticipated Completion Date: 4Q20
C. Anticipated Number of Patients per Site: Centralized recruitment and follow-up
D. Anticipated Number of Sites: Centralized recruitment and follow-up

| Generic Name | Strength and Dosage Form | Therapeutic Classification |
|---|---|---|
| Hesperidin | 500 mg capsule | Antiviral and immune-modulatory natural product |
| | Placebo to match 500 mg capsule | NA |

### 1 INTRODUCTION

Emerging viruses are a major threat to human health and the rate at which new pathogenic viruses are emerging has accelerated in the past fifty years. Generally, antiviral drug development has focused on targeting viral components to block virus entry, replication, morphogenesis and propagation, or on the modulation of the host immune response. The frequent outbreaks have highlighted the urgent need for new antiviral treatments.

#### 1.1 Background

COVID-19 disease

COVID-19 is due to an infection by the beta-coronavirus SARS-CoV-2 (1). The outbreak of COVID-19 disease started in December 2019 in Wuhan, Hubei Province, China (1,2), has spread to other parts of China and Asia, and is now a pandemic that has reached Europe and North America. The number of confirmed cases has reached over 12 507 849, including 560 460 deaths, as of July 12, 2020. Patients initially present with fever with or without respiratory symptoms, but a large number of patients later develop various degrees of pulmonary abnormalities on chest imaging (3). Although the vast majority of patients only have a common, mild form of illness, approximately 15% of the patients fall into the severe group, with requirement of assisted ventilation and oxygenation (3). These patients suffer from acute respiratory distress syndrome. Pathologic findings reveal edema and prominent proteinaceous exudates, vascular congestion, and inflammatory clusters with fibrinoid material and multinucleated giant cells (4).

This single-strand RNA virus depends on angiotensin converting enzyme 2 (ACE2) for its entry into cells (5). ACE2 is primarily produced in Clara cells and type II alveolar epithelial cells (6).

Accumulating evidence suggests that a subgroup of patients with severe COVID-19 might have also a cytokine storm syndrome (7). By blocking virus entry as well as reducing this dysregulated (exaggerated) inflammatory response has been advocated to address the immediate need to reduce the rising mortality (7,8). Therefore, we hypothesize that Hesperidin, by targeting virus entry (binding to ACE2) and by reducing inflammation, may reduce COVID-19 infection and complications in adults at risk or with evidence of acute respiratory distress syndrome.

#### 1.2 Background on Hesperidin

Hesperidin, a flavonoid naturally present in citrus fruits, is a heteroside of a sugar (rutinose) and hesperetin. Hesperidin was discovered in 1828 and is the oldest commercially available flavonoid produced from citrus fruits. Hesperetin inhibits the enzyme 3CL protease that contributes to SARS-CoV-2 replication (14). Pharmacological modeling further suggests that Hesperetin could interfere with the regional binding domain (RBD) between the spike protein of COVID-19 and its receptor on human endothelial and epithelial cells (ACE2) (15). Furthermore, Hesperidin/Hesperetin has also been shown to have anti-inflammatory effects and to reduce acute lung injury in models of acute respiratory distress syndromes induced by Lipopolysaccharides (LPS) and mechanical injury (16-19). Hesperidin could therefore interfere with three operative pathways of COVID-19 infection: SARS-CoV-2 binding to its lung epithelial cell receptor ACE2, reduction of viral replication by inhibition of 3CL protease and modulation of excessive lung inflammation and injury leading to respiratory failure.

Hesperidin is already readily available in consumer products such as orange juice (150-200 mg per serving) and natural products from citrus extracts (up to 500 mg per dose). These natural products are advertised as “improving capillary fragility”. Hesperidin is also approved by the FDA and EMA as pharmaceutical, food and beverage additive used to provide texture, improve taste and conservation due to its anti-oxidant properties. The FDA issued a Generally Recognized As Safe (GRAS) notice for orange extracts with 85% Hesperidin content to be used in food (GRA No 796). In human, plasma concentration of hesperetin, the active component of hesperidin, reach ups to 2 µM after consumption of 150 mg hesperidin with peaks after 3 hours and with a half-life of 7 hours. Based on safety, preclinical studies, human PK studies, and PK modeling of hesperidin, a dose of 1g BID of hesperidin is recommended for anticipated efficacy (inhibition of SARS-CoV-1 3CLPro: 8 µM). SARS-CoV-1 has 95% homology with SARS-CoV-2 (21). A number of human studies have been performed with hesperidin and are summarized in the FDA GRAS notice of 2018 (GRA No 796). There have been no adverse events observed with high dose hesperidin consumption for long periods of time in human subjects. In particular, Demonty et al. studied 204 subjects and found no adverse events after administration of 800 mg daily hesperidin or placebo in capsules for a duration of 4 weeks (22). Hesperidin is accepted for human use alone or as a component of natural products and has been used for centuries in traditional Chinese medicine as it is major product contained in Chenpi (sun-dried tangerine peels).

##### 1.3 COVID-19 symptoms

The epidemiological and clinical characteristics of COVID-19 infection were reviewed by the “Institut National de Santé Publique du Québec” and are available in an online document. Based on a metanalysis of 43 studies including 3600 subjects the most frequently reported symptoms were fever in 83.3% (95% CI: 78.4 – 87.7), cough in 60.3% (54.2 -2 – 66.3), fatigue in 38% (29.8 – 46.5%). More recently, studies have reported that a loss of smell (anosmia) was present in 59% of subjects for one study in 1702 non-hospitalized COVID-19 positive subjects and in 85.6% of 417 hospitalized subjects in another study. Other less frequent symptoms include chills, sore throat, runny nose, shortness of breath, nausea/vomiting, headache, pain (muscular, chest, abdominal, joint etc.), irritability/confusion and diarrhea. There is therefore great variability in the reported prevalence of COVID-19 symptoms and unknown sensitivity and specificity of any single symptom. For the Hesperidin study, a composite primary endpoint of symptoms was therefore determined based on the current guidelines of the government of Canada for clinicians on COVID-19 symptoms and on guidelines for diagnosis of COVID-19 from the Institut National de la Santé Publique du Québec (INSPQ). The Canadian guidelines for clinicians, reviewed on June 18 2020, list the symptoms in three categories as more frequent (> 50%), less frequent (< 50%) and rare (< 10%) (table 1, available online).

**Table 1. Frequency of COVID-19 symptom according to the Canadian guidelines for clinicians**

| More frequent (>50%) | Less frequent (<50%) | Rare (<10%) |
|---|---|---|
| • Fever (44-91%)<br>• Cough (57-74%)<br>• Shortness of breath (31–63%)<br>• Fatigue (31–70%)<br>• Loss of appetite (39-84%)<br>• Loss of smell and/or taste (54-88%) | • Sputum production (28–33%)<br>• Muscle aches (11–44%)<br>• Chest pain (16-36%)<br>• Diarrhea (5-24%)<br>• Nausea/vomiting (5-19%)<br>• Headache (6-70%)<br>• Dizziness (9-17%)<br>• Sore throat (11-13%) | • Confusion<br>• Runny nose<br>• Fainting<br>• Skin manifestations |
| Note: Symptoms among older adults (65 years of age and older) and those with underlying medical conditions may be atypical or subtle; for instance they may be more likely to present without fever or respiratory symptoms. | | |

The guidelines for the diagnosis of COVID-19 from the INSPQ separated symptoms into group A and group B symptoms. The four group A symptoms are fever (of more than 38º C), cough (recent or exacerbation of chronic cough), shortness of breath and anosmia. These symptoms have less inherent subjectivity in their perception and are classified as more frequent in the Canadian guidelines (table 1). The Hesperidin study will use these group A symptoms as a composite primary endpoint. A list of the COVID-19 symptoms as they will be recorded in patients' diary of the Hesperidin trial is presented in appendix E. Although a proportion of SARS-CoV-2 infections may be asymptomatic, the exact proportion remains currently unknown. In symptomatic subjects, the symptoms can become quite severe causing disability and requiring hospitalization.

#### 2 STUDY OBJECTIVES

- The **primary objective** of the study is to determine the effect of hesperidin therapy on the presence of selected COVID-19 symptoms (fever, cough, shortness of breath or anosmia) at 3, 7, 10, and 14 days following randomization.

- The **secondary objectives** are:
  1- To determine the effect of hesperidin therapy on the mean number of COVID-19 symptoms at 3, 7, 10, and 14 days following randomization.
  2- To determine the effect of hesperidin therapy on the duration of COVID-19 symptoms.
  3- To determine the effect of hesperidin therapy on the presence of each individual COVID-19 symptoms at 3, 7, 10, and 14 days following randomization.

- The **exploratory objectives** are:
  1- To evaluate the effect of hesperidin therapy on the presence of each individual COVID-19 symptoms on a daily basis based on a patient symptoms' diary.
  2- To evaluate the effect of hesperidin therapy on COVID-19-related hospitalization, mechanical ventilation and death in the 14 days following randomization.

### 3 STUDY DESIGN

This will be a randomized, double-blind, placebo-controlled study. The study will include subjects from Quebec diagnosed with COVID-19 infections. Following informed consent, 216 subjects meeting all inclusion and no exclusion criteria will be randomized to receive either hesperidin (1000 mg q.d.) or placebo (1:1 allocation ratio) for 14 days. Investigational drug will be delivered to the patients' homes. In addition, patients will be provided with an oral electronic thermometer as well as a daily symptoms diary (see appendix). Follow-up phone will occur after 3 7, 10, and 14 days following randomization for evaluation of the occurrence of any trial endpoints or other adverse events. Electronic CRF (eCRF) will be completed by the research personnel over the phone with the patients.

#### 3.1 Study Schedule

The schedule of visits for this study is outlined in Table 1. However, a patient may be evaluated at any time for safety concerns.

#### 3.2 Enrollment

Informed consent will be obtained from patients who volunteer to participate in the study prior to the conduct of any study-specific procedures. The patient will be considered "enrolled" into the study at the time an informed consent is provided.

##### 3.3 Screening Evaluations and Randomization

Screening evaluations will include a review of the patient's medical history and assessment of concomitant medications to determine if the patient qualifies for the study. Patients who meet all inclusion criteria and no exclusion criteria will be randomized to receive the study product or placebo. All patients must be randomized within 2 days of the diagnosis of COVID-19 infection.

Women of childbearing potential must have a negative urine pregnancy test result at the time of randomization in order to qualify. Qualifying patients will be randomized to receive placebo or hesperidin 1000 mg/day administered in a blinded manner. Blinded randomization will be performed through a system of envelops. 108 patients will be randomized to active treatment and 108 patients will be randomized to placebo for a total of 216 randomized patients.

##### 3.4 Active Treatment Period

All patients will receive study medication per os (PO) once daily for 14 days. If a dose is missed, it should not be replaced. Throughout the study, patients will undergo phone contacts (at 3, 7, 10 and days to assess for potential study endpoints and other adverse events (AEs). Patients will be dispensed study medication for the 14 days, an electronic thermometer and a daily symptoms diary. At each phone contact, patients will be 1) questioned about COVID-19 symptoms; 2) questioned in a non-specific manner for the occurrence of AEs and any change in concomitant medications; and 3) encouraged to comply with the study protocol including adherence to study medication and completion of the daily symptoms diary.

Please refer to Table 1 and Appendix A for Timetable of Visits and Procedures.

#### Table 1 Timetable of Visits and Procedures

| **Visits** | **1. Screening / randomization** (Phone call) | **2. Contact Point** (Phone) –no window** | **3. Contact Point** (Phone ) (± 1 day) | **4. Contact Point** (Phone) (± 3 days) | **5. Contact Point** (Phone call) (± 3 days) |
|---|---|---|---|---|---|
| **Days** | **0** | **3** | **7** | **10** | **14** |
| Informed consent | X | | | | |
| Demographics | X | | | | |
| Medical/Surgical history | X | | | | |
| Review concomitant medication | X | X | X | X | X |
| Covid-19 symptoms (including date of onset) | X | X | X | X | X |
| Review Inclusion/Exclusion criteria | X | | | | |
| Urinary pregnancy test* | X | | | | |
| Randomization | X | | | | |
| Record potential study endpoints and other AEs | | X | X | X | X |
| Study medication dispensing | X | | | | |
| Study medication compliance | | X | X | X | X |

*For women of childbearing potential
** If day 3 occurs on the weekend, contact point will be done on previous Friday or following Monday, whichever is closer to day 3.

### 4 STUDY POPULATION

#### 4.1 Source and Number of Patients

A total of 216 patients will be randomized.

#### 4.2 Patient Selection Criteria

##### 4.2.1 Inclusion Criteria:

All of these criteria must be met:

- Covid-19 positive by polymerase chain reaction (PCR) testing;
- Participant must be able to evaluate their symptoms and report them in the symptoms' diary;
- Patients must be able to take their oral temperature daily with an electronic thermometer provided to them with study materials;
- Males and females, at least 18 years of age, capable and willing to provide informed consent;
- Female patient is either not of childbearing potential, defined as postmenopausal for at least 1 year or surgically sterile, or is of childbearing potential and practicing at least one method of contraception and preferably two complementary forms of contraception including a barrier method (e.g. male or female condoms, spermicides, sponges, foams, jellies, diaphragm, intrauterine device (IUD)) throughout the study;
- Patient must have received a diagnosis of COVID-19 infection within the last 48 hours and have one or more symptoms;
- Outpatient setting (not currently hospitalized or under immediate consideration for hospitalization);
- Patient must be able and willing to comply with the requirements of this study protocol.

##### 4.2.2 Exclusion Criteria:

None of these exclusion criteria should be met:

- Patient currently hospitalized or under immediate consideration for hospitalization;
- Patient currently in shock or with hemodynamic instability;

- Patient undergoing chemotherapy for cancer;
- Patient is unable to take oral temperature using an electronic thermometer;
- Patient who received at least one dose of the COVID-19 vaccine;
- Female patient who is pregnant or breast-feeding or is considering becoming pregnant during the study;
- People taking anticoagulant/antiplatelet medications, those with bleeding disorders, and people two weeks before or after surgery;
- Patient is considered by the investigator, for any reason, to be an unsuitable candidate for the study;
- Regular consumption of natural products containing more than 150 mg of hesperidin or regular consumption of more than 1 glass of orange juice per day;
- Known allergy to any of the medicinal and non-medicinal ingredient: hesperidin, microcrystalline cellulose, magnesium stearate.

##### 4.3 Prohibited, Allowable and Concurrent Medications

Clinical research suggests that hesperidin may affect blood clotting and increase the risk of bleeding. Consequently, anticoagulant and antiplatelet medications are not allowed during the study. Other medications are allowed as long as they are stabilized prior to study entry and maintained as stable throughout the course of the trial.

The use of concomitant medications at the time of randomization will be recorded in the eCRF (with the exceptions of concomitant medication taken “when necessary” (PRN)).

The use of natural products containing hesperidin and its content (in mg) as well as the quantity of orange juice consumption (in ml) will be obtained. Daily consumption of more than 150 mg of hesperidin daily or of more than one glass of orange juice daily will represent an exclusion criterion. Subjects will be encouraged to avoid hesperidin-containing supplements and orange juice for the duration of the study.

#### 5 STUDY METHODOLOGY

##### 5.1 Efficacy Outcomes

- The **primary endpoint**:
  Proportion of subjects with any of the following COVID-19 symptoms: fever, cough, shortness of breath or anosmia, at day 3, 7, 10 and 14.

- The **secondary endpoints**:
  1- Mean number of COVID-19 symptoms (range 0 – 13) at day 3, 7, 10 and 14.
  2- Duration of COVID-19 symptoms, defined as the number of days between first symptoms and complete disappearance of any symptom.
  3- For each COVID-19 symptom listed in the appendix, proportion of subjects with the symptom at day 3, 7, 10 and 14.

Duration of symptoms will be determined as follows:
- If randomization and study product reception by participant occur the same day, first day of symptoms will be defined as the day of randomization;
- If randomization and study product reception by participant occur on different days, first day of symptoms will be defined as the day of study product reception.

- The **exploratory endpoints**:
  1- For each COVID-19 symptom listed in the appendix, proportion of subjects with the symptom on a daily basis based on patients' diary.
  2- Composite of COVID-19-related hospitalization, mechanic ventilation or death in the 14 days following randomization.

#### 5.2 Safety Monitoring

Drug safety will be assessed by an evaluation of types, frequencies, severities and duration of any reported AEs.

##### 5.2.1 Adverse Event Reporting

Information regarding AEs will be collected at day 3, 7, 10 and 14 after randomization. Any AEs prior to randomization will be recorded in the medical history and kept in the patients' chart.

All SAEs will be recorded on the appropriate eCRF section. Information collected will include the onset, duration, severity, relationship to study drug, and the management as outlined in Appendix C.

###### 5.2.1.1 Serious Adverse Events

Serious Adverse Events (SAE) are those that meet any of the following International Council for Harmonisation (ICH) criteria:

- Is fatal or immediately life-threatening;

- Results in persistent or significant disability/incapacity;
- Requires or prolongs inpatient hospitalization;
- Is a congenital anomaly/birth defect in the offspring of the patient;
- Is a cancer;
- Is an overdose (intentional or accidental);
- Is judged to be medically important.

Medically important events may not be immediately life-threatening or result in death or hospitalization but may jeopardize the patient or may require intervention to prevent one of the outcomes listed in the definition above. Serious Adverse Events are to be reported if they are known to occur within 14 days after randomization.
Medical and scientific judgment should be exercised in deciding whether other AEs meet these criteria and are immediately reportable to the sponsor or designee.

In the event of a serious or life-threatening adverse event, or in the event of death, immediately report the event on the appropriate SAE form in the eCRF.

FOR ANY SAFETY QUESTIONS OR CONCERNS, PLEASE CONTACT:
**MHICC Medical Monitor**

****

If any SAE occurs, the study treatment may be interrupted or discontinued at the Investigator's discretion. If an acute medical emergency occurs, the investigator may break the randomization code at any time if this is required for proper treatment of the patient and inform the person listed above by email.

The MHICC Medical Monitor or his designated representative is responsible to report any Suspected Unexpected Serious Adverse Reaction (SUSAR) to the regulatory authorities and to notify Ingenew Pharma at time of submission. Reports to the Health Authorities must be made within 7 calendar days, (for death and life-threatening events) and within 15 calendar days (for other serious events) after being informed of an SAE by the investigator.

###### 5.2.1.2 Adverse Event Follow-Up

Record all reportable serious and non-serious AEs on the appropriate eCRF including the onset, duration, severity, relationship to the study drug, and ultimate management. All AEs reported during the treatment phase should be recorded and followed until the AE has subsided, or stabilized or until the end of the study, whichever occurs first. The study will be stopped at any time if new knowledge is gained and the risk-benefit ratio is no longer favorable for the participating patients and Ingenew Pharma will then be informed immediately but no later than 3 business days if such decision is made

#### 5.3 Withdrawal of Patients from the Study

Patients have the right to withdraw from the study at any time during the course of the study. However, every effort should be made, within the bounds of safety, patient choice and the provisions of informed consent, to have each patient complete the study up to and including the last protocol-specified phone call at Day 14. If the study medication jeopardizes the patient's health or if the patient wishes to discontinue for any reason, study medication can be discontinued but the patient should be encouraged to remain in the study for the follow-up phone calls up to Day 14. Patients who are not compliant during the active treatment period should be counseled on the importance of complying with study requirements and be allowed to remain in the study. No patient who has withdrawn his/her consent from the study during the active treatment period should be replaced.

In the case of an adverse event or safety concern, the study medication may be withheld temporarily, or the dose reduced, as per investigator judgment. Investigators should discuss the patient condition with the MHICC medical monitor in order to determine the appropriate dose reduction algorithm or study medication discontinuation plan.

Patients withdrawn at any time from the study during the active treatment period should complete all last protocol-specified phone call assessments at Day 14 (the reason for withdrawal from the study, the date of the last videovisit/phone call and the date of the last dose of double-blind medication will be clearly documented in the eCRF).

##### 5.4 Study Completion

The study will end when the last randomized patient will have completed his (or her) 14-day follow-up phone contact or videovisit. Completion of the study by a patient should be clearly indicated in the eCRF, along with the date of the last phone call and the date of the last dose of double-blind medication.

#### 6 STUDY MEDICATION

Ingenew Pharma will provide both hesperidin and placebo in bulk to MHI pharmacists who will prepare the bottles of 32 capsules of 500 mg each. At the time of randomization, the randomization number will be recorded on the appropriate eCRF section. Blinded medication will be provided as 500 mg hesperidin capsules or matching placebo capsules. A detailed set of dispensing instructions will be provided by the MHICC.

##### 6.1 Medication Dispensing

Study medication may be dispensed by the designated pharmacist according to a detailed set of dispensing instructions. Study medication will be shipped directly to the patient's home.

##### 6.2 Dosage Regimen

At randomization, patients will be dispensed hesperidin capsule or matching placebo. Patients will be instructed to take two capsules once daily at bedtime according to the detailed set of dispensing instructions outlined on the label.
Orally, hesperidin is generally well tolerated. Headache, asthenia, muscle cramps, and gastrointestinal side effects such as abdominal pain, dyspepsia, nausea, and diarrhea may occur in some patients. There may be unknown risks with taking the investigational product.

### 7 DATA COLLECTION

Electronic Case Report Forms (eCRF) for all patients will be supplied by MHICC. These are to be completed as instructed.

### 8 DATA ANALYSIS AND STATISTICAL CONSIDERATIONS

#### 8.1 Statistical Power and Sample Size Considerations

Sample size is based on the proportion of subjects with any of the following COVID-19 symptoms: fever, cough, shortness of breath or anosmia at day 7 since this time point will be the primary focus. Using a two-sided 0.05 significance level, sample sizes of 91 in both groups (182 total) will achieve 80% power to detect an absolute between groups difference of 20% in the proportion of subjects with the above COVID-19 symptoms at day 7. This is assuming that the proportions of patients with the above COVID-19 symptoms at day 7 will be 50% in the placebo group and 30% in the active treatment group. Factoring in a 15% drop out rate, a total of 216 patients will be randomized in the study (108 per group).

#### 8.2 Analysis population

##### 8.2.1 Intent to Treat (ITT) population

All patients randomized will be included in the ITT population. Patients will be assigned to treatment groups as randomized for analysis purposes.

##### 8.2.2 Safety population

All patients who received at least one dose of study medication will be included in the safety analysis population. Patients will be assigned according to the true treatment received for analysis purposes.

#### 8.3 Data analysis

##### 8.3.1 Analysis of efficacy outcomes

Efficacy analysis will be based on the intent-to-treat principle. The primary analysis will compare the proportions of subjects with fever, cough, shortness of breath or anosmia between the two treatment groups using a  generalized linear mixed model with terms for treatment group (placebo, hesperidin), time (3, 7, 10 and 14 days) and treatment group x time interaction. The primary analysis will be the comparison between treatment groups at day 7, although treatment groups will also be compared at the other time points. Mean number of COVID-19 symptoms will be compared between treatment groups using a Student t-test or a Wilcoxon rank-sum test according to distribution. If more appropriate, a Poisson regression model will be used. Duration of COVID-19 symptoms will be compared between treatment groups using a log rank test and Kaplan-Meier curves will be presented. Subjects who will still have COVID-19 symptoms at the end of the study will be censored at the day of their last phone contact or videovisit. The analysis of individual COVID-19 symptoms over time will be similar to the analysis of the primary endpoint and will use generalized linear mixed models. Composite of COVID-19-related hospitalization, mechanic ventilation or death will be compared between treatment groups using a chi-square test. All statistical tests will be two-sided and conducted at the 0.05 significance level. Statistical analysis will be done using SAS version 9.4 or higher.

Full details of the statistical analysis will be described in a statistical analysis plan that will be finalized and approved prior to database lock.

##### 8.3.2 Analysis of safety outcomes

Safety of hesperidin will be evaluated by presenting descriptive statistics on adverse events and serious adverse events broken down by group. This will be done for the population of patients who received at least one dose of study medication (safety population).

##### 8.3.3 Interim analysis

A fully independent 3-member Data and Safety Monitoring Board (DSMB) will be established and will review unblinded safety data as detailed in the DSMB charter after randomization of 50 subjects and after randomization of 100 subjects. If necessary, an additional meeting will take place after randomization of 150 subjects. The DSMB charter will pre-specify the rules for early study termination, approved by all board members.

#### 9 STUDY COORDINATION

The MHICC will be responsible for processing and quality control of the data. Project management will be carried out as described in the MHICC standard operating procedures (SOPs) for clinical studies. The handling of data, including data quality control, will comply with all applicable regulatory guidelines, MHICC SOPs and the study Data Management Plan.

#### LIST OF APPENDICES

| Appendix | Title |
|---|---|
| A | Detailed Study Safety Parameters |
| B | Administrative Procedures for the Reporting of Adverse Events |
| C | Other Administrative and Regulatory Procedure |
| D | Declaration of Helsinki |
| E | Symptoms diary |

Appendix A **Detailed Study Safety Parameters**

#### 1. Medical/Surgical History

The following elements of medical/surgical history will be recorded at the Screening Visit:

- age
- ethnic origin
- sex
- smoking history
- history of diabetes
- history of hypertension
- history of dyslipidemia
- prior MI
- prior PCI
- prior CABG
- prior stroke
- prior heart failure
- prior obstructive pulmonary disease
- prior other respiratory disease

#### 2. Physical Appearance

1. weight
2. height
3. waist circumference

Appendix B **Administrative Procedures for the Reporting of Adverse Events**

The following administrative procedures for reporting AEs are to be followed during the conduct of this clinical trial.

#### 1 ADVERSE EVENTS DURING THE TRIAL

Each patient will be observed and queried in a non-specific manner by the investigator or study coordinator at each visit for any new or continuing AE since the previous visit. Any AEs prior to the first dose of study medication will be recorded in the medical history and kept in the patients' chart. All SAEs will be recorded in the appropriate eCRF section. Information collected will include the onset, duration, severity, relationship to study drug, and the management. SAEs are to be reported if they are known to occur within 21 days after randomization.

The investigator will review concomitant medications being taken by the patient.

**Definitions**

##### 1.1 Pre-existing condition

A pre-existing condition is one that is present prior to randomization. A worsening of a pre-existing condition after taking the first dose of investigational product should be reported as an AE.

##### 1.2 Adverse Event (AE)

An AE is defined as any unfavorable and unintended sign (including a clinically meaningful abnormal laboratory finding), symptom, or disease temporally associated with the use of an investigational product, whether or not related to the investigational product.

A medical procedure is not considered and should not be reported as an AE. However, the medical condition which led to the procedure should be considered as an AE and be reported as such.

##### 1.3 Related Adverse Event

A related AE is one where, according to the Investigator, there is a reasonable possibility that the event may have been caused by the study product.

##### 1.4 Serious Adverse Event (SAE)

Serious Adverse Events (SAE) are those that meet any of the following International Council for Harmonisation (ICH) criteria:

- Is fatal or immediately life-threatening (NOTE: the term "Life-Threatening" refers to an event in which the patient was at immediate risk of death at the time of the event; it does not refer to an event which could hypothetically have caused death had it been more severe);
- Results in persistent or significant disability/incapacity;
- Requires or prolongs patient hospitalization;
- Is a congenital anomaly/birth defect in the offspring of the patient;
- Is a cancer;
- Is an overdose (intentional or accidental);
- Is judged to be medically important.

Medically important events may not be immediately life-threatening, result in death or hospitalization, but may jeopardize the patient or may require intervention to prevent one of the outcomes listed in the definition above. Serious adverse events are to be reported if they are known to occur within 21 days after randomization.

Medical and scientific judgment should be exercised in deciding whether other AEs meet these criteria and are immediately reportable to the sponsor or its designee.

Hospitalization is defined as a patient admission to a hospital for medical treatment or observation; a visit to the emergency room for an outpatient consultation is not considered a hospitalization. Moreover, hospitalization for therapy of the target disease(s) of the study if the protocol explicitly anticipated and defined the symptoms or episodes is not considered SAEs.

##### 1.5 Life-Threatening Adverse Event

A life-threatening AE is an AE that, in the opinion of the investigator, places the patient at immediate risk of death from the reaction as it occurred.

##### 1.6 Clinical Laboratory Adverse Event

A clinical laboratory abnormality is regarded as an AE if it has been confirmed by at least 1 repeat test and suggests a disease and/or organ toxicity severe enough to require active management.

##### 1.7 Treatment-Emergent Signs and Symptoms (TESS)

A TESS event is any AE that was not present prior to randomization or that worsens in character, intensity or frequency while the patient is in an active treatment period.

##### 1.8 Post-treatment Adverse Event

A post-treatment AE is any AE that occurs after treatment is discontinued.

### 2 HANDLING OF ADVERSE EVENTS

#### 2.1 Treatment-Emergent Signs and Symptoms

Any condition/diagnosis that meets the definition of a TESS event is captured adverse event from and kept in the patient's chart.

#### 2.2 Serious Adverse Events

All SAEs are to be immediately reported as outlined in Section 5.2.1.1., Serious Adverse Events, within 24 hours of the Investigator's first knowledge of the event.

#### 2.3 Intensity

The following criteria are used to assess the intensity of each AE:

- Mild: The patient is aware of the sign or symptom, but finds it easily tolerated.
- Moderate: The patient has enough discomfort to cause interference with or change in usual activities.
- Severe: The patient is incapacitated and unable to work or participate in many or all usual activities.

#### 2.4 Relationship to Study Drug – Physician's Assessment

Causal relationship between study drug and an AE will be reported by physician's assessment as follows: not related, possibly and probably.

#### 2.5 Clinical Outcome

The following categories are used to assess the clinical outcome of each AE:

- Recovered – The patient has fully recovered from the AE with or without observable residual effects.
- Not Yet Recovered – the patient is still being treated for the residual effects of the original AE. This does not include treatment for pre-existing conditions including the indication for the study drug.
- With sequela.
- Died Due to this Adverse Event
- Died, Other Causes
- Unknown
- Surgery/Procedure

### 3 CAPTURING AVDERSE EVENTS

#### 3.1 Pre-Existing Condition

A pre-existing condition should be captured in the medical history and kept in the patient's chart.

#### 3.2 Clinical Laboratory Adverse Event

A clinical laboratory abnormality should be reported as an AE only if it is considered to be clinically significant by the investigator and confirmed by repeat testing.

#### 3.3 Hospitalization or Surgery/Procedure

Any AE that results in hospitalization (admitted and not just an emergency room visit) should be reported as an SAE. Any condition/diagnosis responsible for surgery/procedure should be reported as an AE if it meets the criteria for an AE. A medical procedure is not considered and should not be reported as an AE. The surgery/procedure itself will be reported as a Clinical Outcome of the underlying event. Events that prolong any hospitalization are reported as SAEs.

#### 3.4 Death

The cause of death should be reported as an AE.

### 4 REPORTING TO THE SPONSOR

#### 4.1 Immediately Reportable Adverse Events

If an AE meets the definition of Serious, it must be reported IMMEDIATELY in the eCRF. If the eCRF is not available, the investigator should send the paper SAE form by fax (+1-514-461-1301) as soon as possible and within 24 hours of knowledge of the event. Upon return of the availability of the eCRF, the information written on the paper SAE form must be recorded in the eCRF. For any questions or concerns, the investigator may send an email to the MHICC Medical Monitor at ****. If any SAE occurs the investigator, at his discretion, can withdraw the patient from the study while taking the appropriate follow-up action.

MHICC Medical Monitor or his designated representative is responsible to report SUSARs to Regulatory Authorities and to notify Ingenew Pharma.

#### 4.2 Other Adverse Events

Study endpoints are to be reported in the eCRF within 5 days of awareness of the event.

In addition to SAEs and study endpoints, the only AEs to be recorded on the eCRF are those that are judged related to the study medication by the investigator or that are laboratory abnormalities judged clinically significant by the investigator. Information collected will include the onset, duration, severity, relationship to study drug, and the management.

##### 4.3 Follow-Up Period

For SAEs, the patient must remain under observation until the SAE has subsided or stabilized and all serious findings have returned to normal or stabilized. Any follow-up information to an initial SAE report must be updated in the eCRF. Serious Adverse Events are to be reported if they are known to occur within 21 days of randomization.

#### Appendix C Other Administrative and Regulatory Procedures

This appendix provides information necessary to administer the study in compliance with global Good Clinical Practice (GCP) and government regulations.

Your signature on this cover page of the protocol, subsequent amendments, addenda, and the Clinical Trial Agreement confirms that:

- You have been given appropriate information on the study drug
- You have read and understand the protocol and appendices
- You agree to conduct the study in accordance with the provisions of the protocol and applicable regulations
- You acknowledge the sponsor's ownership of the data and results obtained from the conduct of the protocol
- You agree to maintain the confidentiality of information as outlined in this protocol

### 1. ADMINISTRATIVE PROCEDURES

#### 1.1 Ethics and Informed Consent

##### 1.1.1 Declaration of Helsinki

This study will be conducted in accordance with the Declaration of Helsinki.

##### 1.1.2 Ethics Committee (EC) Review and Approval of the Study

An EC that is organized and operates according to GCP and applicable laws and regulations, should safeguard the rights, safety, and well-being of all trial patients. No patient should be admitted to a trial before the EC issues its written approval/favorable opinion of the trial.

The investigator is responsible for:

- Promptly reporting to the EC all changes in the research activity, all unlabeled AEs, and all unanticipated problems involving risks to human patients or others;
- Not making any changes in the research without EC approval, except when absolutely necessary to eliminate apparent immediate hazards to human patients;
- Submitting a progress report describing the status of the clinical investigation to the EC at appropriate intervals not exceeding 1 year; and
- Submitting a final report when required by the EC within 3 months following completion, termination or discontinuation of the study. Copies of these reports will also be provided **to the sponsor or the sponsor's designated representative**.

In general, all communications with the EC regarding the study will be handled by the principal investigator or coordinating investigator, (if applicable) of the study. The sponsor or the sponsor's designated representative may directly contact the EC if necessary, but must not attempt to influence the EC in any way. A copy of all communications from the EC to the investigator regarding its review of an initial approval of the study and its re-approvals at intervals must be provided to the sponsor by the investigator.

##### 1.1.3 Informed Consent Form (ICF)

The investigator or research personnel will fully explain the purpose of the study to the patient or his/her guardian prior to entering the patient into the study. Informed consent will be obtained from patients who volunteer to participate in the study prior to the conduct of any study-specific procedures. The patient will be considered "enrolled" into the study at the time an informed consent is provided.

##### 1.1.4 Emergency Code Breaks

The investigator will be provided with a mechanism for emergency determination of a patient's treatment regimen (if required for proper treatment of the patient).

##### 1.1.5 Confidentiality of Patient Information

All patients will be assigned a patient number. Subsequently, patients will be identified in the eCRF only by that number. Any information published as a result of the study will be such that it will not permit identification of any patient. The information from this study will be available within the sponsor organization and may be shared with the regulatory authorities. It may also be the subject of an audit by a regulatory agency within the local government. The patient's identity will remain protected except as required for legal or regulatory inquiries.

##### 1.1.6 Publication of Study Results

All information and data regarding the study drug obtained in connection with the conduct of this study are considered confidential. Accordingly, the sponsor (Montreal Heart Institute) retains the right to review manuscripts, abstracts, and presentation material related to this protocol and its amendments/addenda prior to presentation or submission to a journal. This review will not restrict publication of facts or opinions formulated by the investigator.

##### 1.1.7 Roles and Responsibilities

Montreal Heart Institute (MHI)
The Montreal Heart Institute is responsible for the conduction of the study.

Montreal Heart Institute Foundation:
The Montreal Heart Institute Foundation is the sponsor for this study.

Montreal Health Innovations Coordinating Center (MHICC)

The MHICC is a division of the Montreal Heart Institute, and has been designated to manage and coordinate this trial. These activities include overall project management, site selection, distribution of study materials, site management including clinical monitoring, SUSARs reporting to regulatory authorities, data management, biostatistical analysis and writing of the final clinical study report.

Ingenew Pharma Inc.
Ingenew Pharma Inc. will provide the Montreal Heart Institute with study medication and matched placebo for all patients for the duration of this trial according to a Clinical Supply Agreement between them and MHI.

Investigator
The Investigator is responsible for ensuring this trial is conducted according to the signed investigator statement, following ICH/GCP guidelines and all other local regulatory requirements; for protecting the rights, safety, and welfare of subjects under the investigator's care; and ensure accountability of the investigational product.

It is the responsibility of the investigator(s) that:

- The study is conducted in accordance with the Declaration of Helsinki and according to the guidelines in the attached appendices.
- This study is conducted in compliance with all applicable laws and regulations of the local and country where the study is conducted.
- This study is not initiated until the protocol and a copy of the informed and consent form (ICF) have been reviewed and approved by a duly constituted Ethics Committee (EC), and that any local institutional requirements are satisfied.
- Each patient and/or their legal guardian (or caregiver) reads, understands, and signs an instrument of informed consent.
- The patient be informed that personal information may be examined during audit by properly authorized individuals but that personal information will be treated as strictly confidential and not be publicly available.
- The patient log and patient records are retained as detailed in this protocol.

The final responsibility for the content of the informed consent statement remains with the investigator and the EC. Indemnification of the investigator, coworkers, and the institution is provided as specified in the Clinical Trial Agreement.

#### 2. PROTOCOL AMENDMENTS AND ADDENDA

##### 2.1 Definitions

A protocol amendment is any systematic change (e.g., revision, addition, deletion) that is made to the Final Protocol in a clinical study and is identified by consecutive Arabic numerals (e.g.,

Amendment 1, Amendment 2, etc.). Amendments can be made regardless of whether the protocol has been signed by the investigator or whether or not the protocol has been implemented at a site.

A Protocol Addendum is any systematic change (e.g., revision, addition, deletion) that is made to the Final Protocol and is identified by single sequentially ordered letters (e.g., Addendum A, Addendum B).

An Urgent Protocol Amendment is one that must be instituted quickly, usually to eliminate an apparent immediate hazard to subjects and may be implemented prior to eventual EC review (within 5 working days) and submission to regulatory authorities.

All amendments/addenda to the protocol must be approved by the principal investigator, the sponsor and the EC of the investigator's institution. The investigator is responsible for submitting any proposed change in the approved protocol in writing to the EC for review and approval and for sending a copy of the approval to the sponsor or designee. All amendments/addenda will be filed with appropriate local regulatory authorities by the sponsor or designee.

With the exception of urgent protocol amendments, as outlined below, the amendment/addendum will apply to all subjects/patients entered into the study (or all subjects/patients in affected sites for addenda) after it has gone through the applicable procedure described above and been approved by the EC. Any amendments/addenda proposed in a multicenter protocol must be approved by the EC at the individual study site before it can be placed in effect at that site.

##### 2.2 Urgent Protocol Amendment

If the amendment eliminates an apparent immediate safety hazard to the patient (urgent protocol amendment), it may be implemented immediately. The sponsor will promptly notify the appropriate regulatory authorities of the amendment while the investigator will notify his/her EC of the change in writing within 5 working days of its implementation.

#### 3. STUDY TERMINATION

The study will normally be carried to completion as described in the protocol. However, if in the course of the study a severe adverse reaction or intercurrent illness is noted in any patient, consideration may be given to abrupt termination of the study for this patient. Such a decision may be made by either the principal investigator or by the sponsor, or both. Likewise, the study may be terminated due to ethical/safety issues or at the sponsor’s discretion or for regulatory issues.

**Appendix D Declaration of Helsinki**

Adopted by the 18th WMA General Assembly, Helsinki, Finland, June 1964
and amended by the:
29th WMA General Assembly, Tokyo, Japan, October 1975
35th WMA General Assembly, Venice, Italy, October 1983
41st WMA General Assembly, Hong Kong, September 1989
48th WMA General Assembly, Somerset West, Republic of South Africa, October 1996
52nd WMA General Assembly, Edinburgh, Scotland, October 2000
53rd WMA General Assembly, Washington DC, USA, October 2002 (Note of Clarification added)
55th WMA General Assembly, Tokyo, Japan, October 2004 (Note of Clarification added)
59th WMA General Assembly, Seoul, Republic of Korea, October 2008
64th WMA General Assembly, Fortaleza, Brazil, October 2013

**Preamble**

1. The World Medical Association (WMA) has developed the Declaration of Helsinki as a statement of ethical principles for medical research involving human subjects, including research on identifiable human material and data.

   The Declaration is intended to be read as a whole and each of its constituent paragraphs should be applied with consideration of all other relevant paragraphs.

2. Consistent with the mandate of the WMA, the Declaration is addressed primarily to physicians. The WMA encourages others who are involved in medical research involving human subjects to adopt these principles.

**General Principles**

3. The Declaration of Geneva of the WMA binds the physician with the words, “The health of my patient will be my first consideration,” and the International Code of Medical Ethics declares that, “A physician shall act in the patient's best interest when providing medical care.”

4. It is the duty of the physician to promote and safeguard the health, well-being and rights of patients, including those who are involved in medical research. The physician's knowledge and conscience are dedicated to the fulfillment of this duty.

5. Medical progress is based on research that ultimately must include studies involving human subjects.

6. The primary purpose of medical research involving human subjects is to understand the causes, development and effects of diseases and improve preventive, diagnostic and therapeutic interventions (methods, procedures and treatments). Even the best proven interventions must be evaluated continually through research for their safety, effectiveness, efficiency, accessibility and quality.

7. Medical research is subject to ethical standards that promote and ensure respect for all human subjects and protect their health and rights.

8. While the primary purpose of medical research is to generate new knowledge, this goal can never take precedence over the rights and interests of individual research subjects.

9. It is the duty of physicians who are involved in medical research to protect the life, health, dignity, integrity, right to self-determination, privacy, and confidentiality of personal information of research subjects. The responsibility for the protection of research subjects must always rest with the physician or other health care professionals and never with the research subjects, even though they have given consent.

10. Physicians must consider the ethical, legal and regulatory norms and standards for research involving human subjects in their own countries as well as applicable international norms and standards. No national or international ethical, legal or regulatory requirement should reduce or eliminate any of the protections for research subjects set forth in this Declaration.

11. Medical research should be conducted in a manner that minimizes possible harm to the environment.

12. Medical research involving human subjects must be conducted only by individuals with the appropriate ethics and scientific education, training and qualifications. Research on patients or healthy volunteers requires the supervision of a competent and appropriately qualified physician or other health care professional.

13. Groups that are underrepresented in medical research should be provided appropriate access to participation in research.

14. Physicians who combine medical research with medical care should involve their patients in research only to the extent that this is justified by its potential preventive, diagnostic or therapeutic value and if the physician has good reason to believe that participation in the research study will not adversely affect the health of the patients who serve as research subjects.

15. Appropriate compensation and treatment for subjects who are harmed as a result of participating in research must be ensured.

**Risks, Burdens and Benefits**

16. In medical practice and in medical research, most interventions involve risks and burdens.

    Medical research involving human subjects may only be conducted if the importance of the objective outweighs the risks and burdens to the research subjects.

17. All medical research involving human subjects must be preceded by careful assessment of predictable risks and burdens to the individuals and groups involved in the research in comparison with foreseeable benefits to them and to other individuals or groups affected by the condition under investigation.

    Measures to minimize the risks must be implemented. The risks must be continuously monitored, assessed and documented by the researcher.

18. Physicians may not be involved in a research study involving human subjects unless they are confident that the risks have been adequately assessed and can be satisfactorily managed.

    When the risks are found to outweigh the potential benefits or when there is conclusive proof of definitive outcomes, physicians must assess whether to continue, modify or immediately stop the study.

**Vulnerable Groups and Individuals**

19. Some groups and individuals are particularly vulnerable and may have an increased likelihood of being wronged or of incurring additional harm.

    All vulnerable groups and individuals should receive specifically considered protection.

20. Medical research with a vulnerable group is only justified if the research is responsive to the health needs or priorities of this group and the research cannot be carried out in a non-vulnerable group. In addition, this group should stand to benefit from the knowledge, practices or interventions that result from the research.

**Scientific Requirements and Research Protocols**

21. Medical research involving human subjects must conform to generally accepted scientific principles, be based on a thorough knowledge of the scientific literature, other relevant

sources of information, and adequate laboratory and, as appropriate, animal experimentation. The welfare of animals used for research must be respected.

22. The design and performance of each research study involving human subjects must be clearly described and justified in a research protocol.
    The protocol should contain a statement of the ethical considerations involved and should indicate how the principles in this Declaration have been addressed. The protocol should include information regarding funding, sponsors, institutional affiliations, potential conflicts of interest, incentives for subjects and information regarding provisions for treating and/or compensating subjects who are harmed as a consequence of participation in the research study.

    In clinical trials, the protocol must also describe appropriate arrangements for post-trial provisions.

**Research Ethics Committees**

23. The research protocol must be submitted for consideration, comment, guidance and approval to the concerned research ethics committee before the study begins. This committee must be transparent in its functioning, must be independent of the researcher, the sponsor and any other undue influence and must be duly qualified. It must take into consideration the laws and regulations of the country or countries in which the research is to be performed as well as applicable international norms and standards but these must not be allowed to reduce or eliminate any of the protections for research subjects set forth in this Declaration.

    The committee must have the right to monitor ongoing studies. The researcher must provide monitoring information to the committee, especially information about any serious adverse events. No amendment to the protocol may be made without consideration and approval by the committee. After the end of the study, the researchers must submit a final report to the committee containing a summary of the study's findings and conclusions.

**Privacy and Confidentiality**

24. Every precaution must be taken to protect the privacy of research subjects and the confidentiality of their personal information.

**Informed Consent**

25. Participation by individuals capable of giving informed consent as subjects in medical research must be voluntary. Although it may be appropriate to consult family members or community leaders, no individual capable of giving informed consent may be enrolled in a research study unless he or she freely agrees.

26. In medical research involving human subjects capable of giving informed consent, each potential subject must be adequately informed of the aims, methods, sources of funding, any possible conflicts of interest, institutional affiliations of the researcher, the anticipated benefits and potential risks of the study and the discomfort it may entail, post-study provisions and any other relevant aspects of the study. The potential subject must be informed of the right to refuse to participate in the study or to withdraw consent to participate at any time without reprisal. Special attention should be given to the specific information needs of individual potential subjects as well as to the methods used to deliver the information.

    After ensuring that the potential subject has understood the information, the physician or another appropriately qualified individual must then seek the potential subject's freely-given informed consent, preferably in writing. If the consent cannot be expressed in writing, the non-written consent must be formally documented and witnessed.

    All medical research subjects should be given the option of being informed about the general outcome and results of the study.

27. When seeking informed consent for participation in a research study the physician must be particularly cautious if the potential subject is in a dependent relationship with the physician or may consent under duress. In such situations the informed consent must be sought by an appropriately qualified individual who is completely independent of this relationship.

28. For a potential research subject who is incapable of giving informed consent, the physician must seek informed consent from the legally authorized representative. These individuals must not be included in a research study that has no likelihood of benefit for them unless it is intended to promote the health of the group represented by the potential subject, the research cannot instead be performed with persons capable of providing informed consent, and the research entails only minimal risk and minimal burden.

29. When a potential research subject who is deemed incapable of giving informed consent is able to give assent to decisions about participation in research, the physician must seek that assent in addition to the consent of the legally authorized representative. The potential subject's dissent should be respected.

30. Research involving subjects who are physically or mentally incapable of giving consent, for example, unconscious patients, may be done only if the physical or mental condition that prevents giving informed consent is a necessary characteristic of the research group. In such circumstances the physician must seek informed consent from the legally authorized representative. If no such representative is available and if the research cannot be delayed, the study may proceed without informed consent provided that the specific reasons for involving subjects with a condition that renders them unable to give informed consent have been stated in the research protocol and the study has been approved by a

research ethics committee. Consent to remain in the research must be obtained as soon as possible from the subject or a legally authorized representative.

31. The physician must fully inform the patient which aspects of their care are related to the research. The refusal of a patient to participate in a study or the patient's decision to withdraw from the study must never adversely affect the patient-physician relationship.

32. For medical research using identifiable human material or data, such as research on material or data contained in biobanks or similar repositories, physicians must seek informed consent for its collection, storage and/or reuse. There may be exceptional situations where consent would be impossible or impracticable to obtain for such research. In such situations the research may be done only after consideration and approval of a research ethics committee.

**Use of Placebo**

33. The benefits, risks, burdens and effectiveness of a new intervention must be tested against those of the best proven intervention(s), except in the following circumstances:

Where no proven intervention exists, the use of placebo, or no intervention, is acceptable; or

Where for compelling and scientifically sound methodological reasons the use of any intervention less effective than the best proven one, the use of placebo, or no intervention is necessary to determine the efficacy or safety of an intervention

and the patients who receive any intervention less effective than the best proven one, placebo, or no intervention will not be subject to additional risks of serious or irreversible harm as a result of not receiving the best proven intervention.

Extreme care must be taken to avoid abuse of this option.

**Post-Trial Provisions**

34. In advance of a clinical trial, sponsors, researchers and host country governments should make provisions for post-trial access for all participants who still need an intervention identified as beneficial in the trial. This information must also be disclosed to participants during the informed consent process.

**Research Registration and Publication and Dissemination of Results**

35. Every research study involving human subjects must be registered in a publicly accessible database before recruitment of the first subject.

36. Researchers, authors, sponsors, editors and publishers all have ethical obligations with regard to the publication and dissemination of the results of research. Researchers have a duty to make publicly available the results of their research on human subjects and are accountable for the completeness and accuracy of their reports. All parties should adhere to accepted guidelines for ethical reporting. Negative and inconclusive as well as positive results must be published or otherwise made publicly available. Sources of funding, institutional affiliations and conflicts of interest must be declared in the publication. Reports of research not in accordance with the principles of this Declaration should not be accepted for publication.

**Unproven Interventions in Clinical Practice**

37. In the treatment of an individual patient, where proven interventions do not exist or other known interventions have been ineffective, the physician, after seeking expert advice, with informed consent from the patient or a legally authorized representative, may use an unproven intervention if in the physician's judgement it offers hope of saving life, re-establishing health or alleviating suffering. This intervention should subsequently be made the object of research, designed to evaluate its safety and efficacy. In all cases, new information must be recorded and, where appropriate, made publicly available.

**Appendix E**

### Symptoms Diary

**Protocol: MHICC-2020-003**

Home self-monitoring on COVID-19 symptoms: The Hesperidin Coronavirus study (Hesperidin)

Patient #: ____ ____ ____ ____ ____

| Diary# | 1 | 2 | 3 | 4 | (*Circle*) |
|---|---|---|---|---|---|

#### Important

Please use the diaries in the order in which they are numbered (1, 2, 3 and 4) and have your complete diary on hand for each phone call.

#### Instructions

This diary is to be used to collect information on the daily symptoms you experience.

This daily diary has been designed with check boxes **to make it easier for you to fill out every day**.

Please check your symptoms every day for 14 days. There is one page for each week that you will be taking the medication starting with Day 1.
We will ask you for the symptoms when we call at the beginning of the study and on day 3, day 7, day 10 and 14. Please have the diary with you.

### Covid-19_ Symptoms Log

**Protocol: MHICC-2020-003**

| **Symptoms** | **Day 1** Yes No | **Day 2** Yes No | **Day 3** Yes No | **Day 4** Yes No | **Day 5** Yes No | **Day 6** Yes No | **Day 7** Yes No |
|---|---|---|---|---|---|---|---|
| Recent cough or aggravation of chronic cough | ☐ ☐ | ☐ ☐ | ☐ ☐ | ☐ ☐ | ☐ ☐ | ☐ ☐ | ☐ ☐ |
| Fever (enter temperature) | | | | | | | |
| Feverish/chills (temperature not taken) | ☐ ☐ | ☐ ☐ | ☐ ☐ | ☐ ☐ | ☐ ☐ | ☐ ☐ | ☐ ☐ |
| Sore throat | ☐ ☐ | ☐ ☐ | ☐ ☐ | ☐ ☐ | ☐ ☐ | ☐ ☐ | ☐ ☐ |
| Runny nose | ☐ ☐ | ☐ ☐ | ☐ ☐ | ☐ ☐ | ☐ ☐ | ☐ ☐ | ☐ ☐ |
| Shortness of breath/difficulty breathing | ☐ ☐ | ☐ ☐ | ☐ ☐ | ☐ ☐ | ☐ ☐ | ☐ ☐ | ☐ ☐ |
| Nausea/vomiting | ☐ ☐ | ☐ ☐ | ☐ ☐ | ☐ ☐ | ☐ ☐ | ☐ ☐ | ☐ ☐ |
| Headache | ☐ ☐ | ☐ ☐ | ☐ ☐ | ☐ ☐ | ☐ ☐ | ☐ ☐ | ☐ ☐ |
| General weakness | ☐ ☐ | ☐ ☐ | ☐ ☐ | ☐ ☐ | ☐ ☐ | ☐ ☐ | ☐ ☐ |
| Pain (muscular, chest, abdominal, joint, etc.) | ☐ ☐ | ☐ ☐ | ☐ ☐ | ☐ ☐ | ☐ ☐ | ☐ ☐ | ☐ ☐ |
| Irritability/confusion | ☐ ☐ | ☐ ☐ | ☐ ☐ | ☐ ☐ | ☐ ☐ | ☐ ☐ | ☐ ☐ |
| Diarrhea | ☐ ☐ | ☐ ☐ | ☐ ☐ | ☐ ☐ | ☐ ☐ | ☐ ☐ | ☐ ☐ |
| Sudden Loss of smell (anosmia) | ☐ ☐ | ☐ ☐ | ☐ ☐ | ☐ ☐ | ☐ ☐ | ☐ ☐ | ☐ ☐ |
| Comment or other, specify: | | | | | | | |

**Reminder: Please have the completed diary on hand at each phone call or video contact follow up.**

| Symptoms | Day 8 | | Day 9 | | Day 10 | | Day 11 | | Day 12 | | Day 13 | | Day 14 | |
|---|---|---|---|---|---|---|---|---|---|---|---|---|---|---|
| | Yes | No | Yes | No | Yes | No | Yes | No | Yes | No | Yes | No | Yes | No |
| Recent cough or aggravation of chronic cough | ☐ | ☐ | ☐ | ☐ | ☐ | ☐ | ☐ | ☐ | ☐ | ☐ | ☐ | ☐ | ☐ | ☐ |
| Fever (enter temperature) | | | | | | | | | | | | | | |
| Feverish/chills (temperature not taken) | ☐ | ☐ | ☐ | ☐ | ☐ | ☐ | ☐ | ☐ | ☐ | ☐ | ☐ | ☐ | ☐ | ☐ |
| Sore throat | ☐ | ☐ | ☐ | ☐ | ☐ | ☐ | ☐ | ☐ | ☐ | ☐ | ☐ | ☐ | ☐ | ☐ |
| Runny nose | ☐ | ☐ | ☐ | ☐ | ☐ | ☐ | ☐ | ☐ | ☐ | ☐ | ☐ | ☐ | ☐ | ☐ |
| Shortness of breath/difficulty breathing | ☐ | ☐ | ☐ | ☐ | ☐ | ☐ | ☐ | ☐ | ☐ | ☐ | ☐ | ☐ | ☐ | ☐ |
| Nausea/vomiting | ☐ | ☐ | ☐ | ☐ | ☐ | ☐ | ☐ | ☐ | ☐ | ☐ | ☐ | ☐ | ☐ | ☐ |
| Headache | ☐ | ☐ | ☐ | ☐ | ☐ | ☐ | ☐ | ☐ | ☐ | ☐ | ☐ | ☐ | ☐ | ☐ |
| General weakness | ☐ | ☐ | ☐ | ☐ | ☐ | ☐ | ☐ | ☐ | ☐ | ☐ | ☐ | ☐ | ☐ | ☐ |
| Pain (muscular, chest, abdominal, joint, etc.) | ☐ | ☐ | ☐ | ☐ | ☐ | ☐ | ☐ | ☐ | ☐ | ☐ | ☐ | ☐ | ☐ | ☐ |
| Irritability/confusion | ☐ | ☐ | ☐ | ☐ | ☐ | ☐ | ☐ | ☐ | ☐ | ☐ | ☐ | ☐ | ☐ | ☐ |
| Diarrhea | ☐ | ☐ | ☐ | ☐ | ☐ | ☐ | ☐ | ☐ | ☐ | ☐ | ☐ | ☐ | ☐ | ☐ |
| Sudden Loss of smell (anosmia) | ☐ | ☐ | ☐ | ☐ | ☐ | ☐ | ☐ | ☐ | ☐ | ☐ | ☐ | ☐ | ☐ | ☐ |
| Comment or other, specify: | | | | | | | | | | | | | | |

**Reminder: Please have the completed diary on hand at each phone call or video contact follow up.**
