## Supplementary material for "Fourteen-days Evolution of COVID-19 Symptoms During the Third Wave in Non-vaccinated Subjects and Effects of Hesperidin Therapy: A randomized, double-blinded, placebo-controlled study": S2 Table

**S2 Table. Proportion of patients with group A COVID-19 symptoms at day 3, 7, 10 and 14 – Attrition bias, intent-to-treat population.**

| **Worst case : Imputation using last observation carried forward when daily symptoms diary was completely missing** | | | | | |
| --- | --- | --- | --- | --- | --- |
|  | **Placebo** | **Hesperidin** | **All** | **OR (95% CI)^a^** | **P-value^a^** |
| **Day 3** | N = 109 | N = 107 | N = 216 |  |  |
| **No** | 13 (11.9%) | 10 (9.3%) | 23 (10.6%) |  |  |
| **Yes^b^** | 96 (88.1%) | 97 (90.7%) | 193 (89.4%) | 1.31 (0.55; 3.16) | 0.5417 |
| **Day 7** | N = 108 | N = 107 | N = 215 |  |  |
| **No** | 25 (23.1%) | 23 (21.5%) | 48 (22.3%) |  |  |
| **Yes^b^** | 83 (76.9%) | 84 (78.5%) | 167 (77.7%) | 1.10 (0.58; 2.10) | 0.7722 |
| **Day 10** | N = 108 | N = 107 | N = 215 |  |  |
| **No** | 40 (37.0%) | 38 (35.5%) | 78 (36.3%) |  |  |
| **Yes^b^** | 68 (63.0%) | 69 (64.5%) | 137 (63.7%) | 1.07 (0.61; 1.87) | 0.8173 |
| **Day 14** | N = 108 | N = 107 | N = 215 |  |  |
| **No** | 44 (40.7%) | 51 (47.7%) | 95 (44.2%) |  |  |
| **Yes^b^** | 64 (59.3%) | 56 (52.3%) | 120 (55.8%) | 0.75 (0.44; 1.30) | 0.3098 |
| **Best case : Imputation assuming no symptom when daily symptoms diary was completely missing** | | | | | |
|  | **Placebo** | **Hesperidin** | **All** | **OR (95% CI)^a^** | **P-value^b^** |
| **Day 3** | N = 109 | N = 107 | N = 216 |  |  |
| **No** | 19 (17.4%) | 14 (13.1%) | 33 (15.3%) |  |  |
| **Yes^b^** | 90 (82.6%) | 93 (86.9%) | 183 (84.7%) | 1.40 (0.66; 2.98) | 0.3786 |
| **Day 7** | N = 108 | N = 107 | N = 215 |  |  |
| **No** | 32 (29.6%) | 33 (30.8%) | 65 (30.2%) |  |  |
| **Yes^b^** | 76 (70.4%) | 74 (69.2%) | 150 (69.8%) | 0.94 (0.53; 1.70) | 0.8474 |
| **Day 10** | N = 108 | N = 107 | N = 215 |  |  |
| **No** | 48 (44.4%) | 49 (45.8%) | 97 (45.1%) |  |  |
| **Yes^b^** | 60 (55.6%) | 58 (54.2%) | 118 (54.9%) | 0.95 (0.55; 1.63) | 0.8431 |
| **Day 14** | N = 108 | N = 107 | N = 215 |  |  |
| **No** | 53 (49.1%) | 68 (63.6%) | 121 (56.3%) |  |  |
| **Yes^b^** | 55 (50.9%) | 39 (36.4%) | 94 (43.7%) | 0.55 (0.32; 0.96) | 0.0343 |

^a^Comparison between placebo group and hesperidin group. Significant when p<0.05

^b^Subject has at least one of the group A COVID-19 symptoms: Fever, cough, shortness of breath or anosmia

N represents the number of subjects who completed the daily symptom diary.
