## Supplementary material for "Fourteen-days Evolution of COVID-19 Symptoms During the Third Wave in Non-vaccinated Subjects and Effects of Hesperidin Therapy: A randomized, double-blinded, placebo-controlled study": S3 Table

2 - Efficacy Analysis
2.2 - Secondary Analysis
Table 2.2.3a - Summary of and Statistical Analysis for Presence of Cough at Day 1 to 14, ITT Population

| | | Placebo N=109 | Hesperidin N=107 | All N=216 |
|---|---|---|---|---|
| DAY 1 | n | 107 | 105 | 212 |
| | No | 40 ( 37.4%) | 43 ( 41.0%) | 83 ( 39.2%) |
| | Yes | 67 ( 62.6%) | 62 ( 59.0%) | 129 ( 60.8%) |
| DAY 2 | n | 106 | 105 | 211 |
| | No | 44 ( 41.5%) | 43 ( 41.0%) | 87 ( 41.2%) |
| | Yes | 62 ( 58.5%) | 62 ( 59.0%) | 124 ( 58.8%) |
| DAY 3 | n | 103 | 102 | 205 |
| | No | 49 ( 47.6%) | 48 ( 47.1%) | 97 ( 47.3%) |
| | Yes | 54 ( 52.4%) | 54 ( 52.9%) | 108 ( 52.7%) |
| DAY 4 | n | 103 | 96 | 199 |
| | No | 46 ( 44.7%) | 47 ( 49.0%) | 93 ( 46.7%) |
| | Yes | 57 ( 55.3%) | 49 ( 51.0%) | 106 ( 53.3%) |
| DAY 5 | n | 102 | 95 | 197 |
| | No | 53 ( 52.0%) | 52 ( 54.7%) | 105 ( 53.3%) |
| | Yes | 49 ( 48.0%) | 43 ( 45.3%) | 92 ( 46.7%) |
| DAY 6 | n | 103 | 92 | 195 |
| | No | 55 ( 53.4%) | 54 ( 58.7%) | 109 ( 55.9%) |
| | Yes | 48 ( 46.6%) | 38 ( 41.3%) | 86 ( 44.1%) |
| DAY 7 | n | 102 | 91 | 193 |
| | No | 57 ( 55.9%) | 54 ( 59.3%) | 111 ( 57.5%) |
| | Yes | 45 ( 44.1%) | 37 ( 40.7%) | 82 ( 42.5%) |
| DAY 8 | n | 101 | 90 | 191 |
| | No | 64 ( 63.4%) | 58 ( 64.4%) | 122 ( 63.9%) |
| | Yes | 37 ( 36.6%) | 32 ( 35.6%) | 69 ( 36.1%) |
| DAY 9 | n | 100 | 89 | 189 |
| | No | 61 ( 61.0%) | 62 ( 69.7%) | 123 ( 65.1%) |
| | Yes | 39 ( 39.0%) | 27 ( 30.3%) | 66 ( 34.9%) |

Note:
Comparaisons at day 3,7,10 and 14 are considered for secondary analysis. The other days are used for exploratory analysis.


CONFIDENTIALITY STATEMENT: Information contained in this statistical report is confidential and should not be disclosed to other parties than those directly involved with the execution of this study without written authorization from Montreal Heart Institute

2 - Efficacy Analysis
2.2 - Secondary Analysis
Table 2.2.3a - Summary of and Statistical Analysis for Presence of Cough at Day 1 to 14, ITT Population

| | | Placebo N=109 | Hesperidin N=107 | All N=216 |
|---|---|---|---|---|
| DAY 10 | n | 100 | 90 | 190 |
| | No | 65 ( 65.0%) | 64 ( 71.1%) | 129 ( 67.9%) |
| | Yes | 35 ( 35.0%) | 26 ( 28.9%) | 61 ( 32.1%) |
| DAY 11 | n | 98 | 86 | 184 |
| | No | 66 ( 67.3%) | 64 ( 74.4%) | 130 ( 70.7%) |
| | Yes | 32 ( 32.7%) | 22 ( 25.6%) | 54 ( 29.3%) |
| DAY 12 | n | 99 | 85 | 184 |
| | No | 68 ( 68.7%) | 62 ( 72.9%) | 130 ( 70.7%) |
| | Yes | 31 ( 31.3%) | 23 ( 27.1%) | 54 ( 29.3%) |
| DAY 13 | n | 98 | 83 | 181 |
| | No | 67 ( 68.4%) | 64 ( 77.1%) | 131 ( 72.4%) |
| | Yes | 31 ( 31.6%) | 19 ( 22.9%) | 50 ( 27.6%) |
| DAY 14 | n | 95 | 79 | 174 |
| | No | 66 ( 69.5%) | 58 ( 73.4%) | 124 ( 71.3%) |
| | Yes | 29 ( 30.5%) | 21 ( 26.6%) | 50 ( 28.7%) |

Note:
Comparaisons at day 3,7,10 and 14 are considered for secondary analysis. The other days are used for exploratory analysis.


CONFIDENTIALITY STATEMENT: Information contained in this statistical report is confidential and should not be disclosed to other parties than those directly involved with the execution of this study without written authorization from Montreal Heart Institute

2 – Efficacy Analysis
2.2 – Secondary Analysis
Table 2.2.3a - Summary of and Statistical Analysis for Presence of Cough at Day 1 to 14, ITT Population (continued)

Generalized linear mixed model (repeated binary logistic regression) with terms for treatment group, time and treatment group x time interaction

| Contrast | OR (95% CI) | P-value |
|---|---|---|
| Hesperidin vs Placebo at Day 3 | 1.02 (0.59; 1.77) | 0.9416 |
| Hesperidin vs Placebo at Day 7 | 0.87 (0.49; 1.55) | 0.6296 |
| Hesperidin vs Placebo at Day 10 | 0.75 (0.41; 1.40) | 0.3711 |
| Hesperidin vs Placebo at Day 14 | 0.82 (0.42; 1.61) | 0.5696 |


CONFIDENTIALITY STATEMENT: Information contained in this statistical report is confidential and should not be disclosed to other parties than those directly involved with the execution of this study without written authorization from Montreal Heart Institute

2 - Efficacy Analysis
2.2 - Secondary Analysis
Table 2.2.3b - Summary of and Statistical Analysis for Presence of Fever at Day 1 to 14, ITT Population

| | | Placebo N=109 | Hesperidin N=107 | All N=216 |
|---|---|---|---|---|
| DAY 1 | n | 105 | 102 | 207 |
| | No | 100 ( 95.2%) | 99 ( 97.1%) | 199 ( 96.1%) |
| | Yes | 5 ( 4.8%) | 3 ( 2.9%) | 8 ( 3.9%) |
| DAY 2 | n | 105 | 103 | 208 |
| | No | 101 ( 96.2%) | 100 ( 97.1%) | 201 ( 96.6%) |
| | Yes | 4 ( 3.8%) | 3 ( 2.9%) | 7 ( 3.4%) |
| DAY 3 | n | 102 | 101 | 203 |
| | No | 96 ( 94.1%) | 96 ( 95.0%) | 192 ( 94.6%) |
| | Yes | 6 ( 5.9%) | 5 ( 5.0%) | 11 ( 5.4%) |
| DAY 4 | n | 102 | 95 | 197 |
| | No | 99 ( 97.1%) | 92 ( 96.8%) | 191 ( 97.0%) |
| | Yes | 3 ( 2.9%) | 3 ( 3.2%) | 6 ( 3.0%) |
| DAY 5 | n | 102 | 94 | 196 |
| | No | 98 ( 96.1%) | 89 ( 94.7%) | 187 ( 95.4%) |
| | Yes | 4 ( 3.9%) | 5 ( 5.3%) | 9 ( 4.6%) |
| DAY 6 | n | 102 | 91 | 193 |
| | No | 99 ( 97.1%) | 88 ( 96.7%) | 187 ( 96.9%) |
| | Yes | 3 ( 2.9%) | 3 ( 3.3%) | 6 ( 3.1%) |
| DAY 7 | n | 101 | 90 | 191 |
| | No | 97 ( 96.0%) | 87 ( 96.7%) | 184 ( 96.3%) |
| | Yes | 4 ( 4.0%) | 3 ( 3.3%) | 7 ( 3.7%) |
| DAY 8 | n | 100 | 90 | 190 |
| | No | 97 ( 97.0%) | 87 ( 96.7%) | 184 ( 96.8%) |
| | Yes | 3 ( 3.0%) | 3 ( 3.3%) | 6 ( 3.2%) |
| DAY 9 | n | 99 | 89 | 188 |
| | No | 98 ( 99.0%) | 87 ( 97.8%) | 185 ( 98.4%) |
| | Yes | 1 ( 1.0%) | 2 ( 2.2%) | 3 ( 1.6%) |

Note:
Comparaisons at day 3,7,10 and 14 are considered for secondary analysis. The other days are used for exploratory analysis.


CONFIDENTIALITY STATEMENT: Information contained in this statistical report is confidential and should not be disclosed to other parties than those directly involved with the execution of this study without written authorization from Montreal Heart Institute

2 - Efficacy Analysis
2.2 - Secondary Analysis
Table 2.2.3b - Summary of and Statistical Analysis for Presence of Fever at Day 1 to 14, ITT Population

| | | Placebo N=109 | Hesperidin N=107 | All N=216 |
|---|---|---|---|---|
| DAY 10 | n | 99 | 90 | 189 |
| | No | 97 ( 98.0%) | 87 ( 96.7%) | 184 ( 97.4%) |
| | Yes | 2 ( 2.0%) | 3 ( 3.3%) | 5 ( 2.6%) |
| DAY 11 | n | 97 | 86 | 183 |
| | No | 96 ( 99.0%) | 84 ( 97.7%) | 180 ( 98.4%) |
| | Yes | 1 ( 1.0%) | 2 ( 2.3%) | 3 ( 1.6%) |
| DAY 12 | n | 98 | 85 | 183 |
| | No | 97 ( 99.0%) | 84 ( 98.8%) | 181 ( 98.9%) |
| | Yes | 1 ( 1.0%) | 1 ( 1.2%) | 2 ( 1.1%) |
| DAY 13 | n | 97 | 83 | 180 |
| | No | 96 ( 99.0%) | 82 ( 98.8%) | 178 ( 98.9%) |
| | Yes | 1 ( 1.0%) | 1 ( 1.2%) | 2 ( 1.1%) |
| DAY 14 | n | 94 | 79 | 173 |
| | No | 93 ( 98.9%) | 79 (100.0%) | 172 ( 99.4%) |
| | Yes | 1 ( 1.1%) | 0 ( 0.0%) | 1 ( 0.6%) |

Note:
Comparaisons at day 3,7,10 and 14 are considered for secondary analysis. The other days are used for exploratory analysis.


CONFIDENTIALITY STATEMENT: Information contained in this statistical report is confidential and should not be disclosed to other parties than those directly involved with the execution of this study without written authorization from Montreal Heart Institute

2 – Efficacy Analysis
2.2 – Secondary Analysis
Table 2.2.3b - Summary of and Statistical Analysis for Presence of Fever at Day 1 to 14, ITT Population (continued)

Generalized linear mixed model (repeated binary logistic regression) with terms for treatment group, time and treatment group x time interaction

| Contrast | OR (95% CI) | P-value |
|---|---|---|
| Hesperidin vs Placebo at Day 3 | 0.83 (0.26; 2.67) | 0.7583 |
| Hesperidin vs Placebo at Day 7 | 0.84 (0.20; 3.58) | 0.8091 |
| Hesperidin vs Placebo at Day 10 | 1.67 (0.30; 9.42) | 0.5591 |
| Hesperidin vs Placebo at Day 14 | 0.00 (0.00; ND) | 0.9983 |


CONFIDENTIALITY STATEMENT: Information contained in this statistical report is confidential and should not be disclosed to other parties than those directly involved with the execution of this study without written authorization from Montreal Heart Institute

2 - Efficacy Analysis
2.2 - Secondary Analysis
Table 2.2.3c - Summary of and Statistical Analysis for Presence of Feverish or Chills at Day 1 to 14, ITT population

| | | Placebo N=109 | Hesperidin N=107 | All N=216 |
|---|---|---|---|---|
| DAY 1 | n | 107 | 105 | 212 |
| | No | 75 ( 70.1%) | 69 ( 65.7%) | 144 ( 67.9%) |
| | Yes | 32 ( 29.9%) | 36 ( 34.3%) | 68 ( 32.1%) |
| DAY 2 | n | 106 | 105 | 211 |
| | No | 82 ( 77.4%) | 70 ( 66.7%) | 152 ( 72.0%) |
| | Yes | 24 ( 22.6%) | 35 ( 33.3%) | 59 ( 28.0%) |
| DAY 3 | n | 103 | 102 | 205 |
| | No | 82 ( 79.6%) | 78 ( 76.5%) | 160 ( 78.0%) |
| | Yes | 21 ( 20.4%) | 24 ( 23.5%) | 45 ( 22.0%) |
| DAY 4 | n | 103 | 96 | 199 |
| | No | 89 ( 86.4%) | 77 ( 80.2%) | 166 ( 83.4%) |
| | Yes | 14 ( 13.6%) | 19 ( 19.8%) | 33 ( 16.6%) |
| DAY 5 | n | 102 | 95 | 197 |
| | No | 89 ( 87.3%) | 79 ( 83.2%) | 168 ( 85.3%) |
| | Yes | 13 ( 12.7%) | 16 ( 16.8%) | 29 ( 14.7%) |
| DAY 6 | n | 103 | 92 | 195 |
| | No | 93 ( 90.3%) | 79 ( 85.9%) | 172 ( 88.2%) |
| | Yes | 10 ( 9.7%) | 13 ( 14.1%) | 23 ( 11.8%) |
| DAY 7 | n | 102 | 91 | 193 |
| | No | 92 ( 90.2%) | 81 ( 89.0%) | 173 ( 89.6%) |
| | Yes | 10 ( 9.8%) | 10 ( 11.0%) | 20 ( 10.4%) |
| DAY 8 | n | 101 | 90 | 191 |
| | No | 91 ( 90.1%) | 82 ( 91.1%) | 173 ( 90.6%) |
| | Yes | 10 ( 9.9%) | 8 ( 8.9%) | 18 ( 9.4%) |
| DAY 9 | n | 100 | 89 | 189 |
| | No | 93 ( 93.0%) | 85 ( 95.5%) | 178 ( 94.2%) |
| | Yes | 7 ( 7.0%) | 4 ( 4.5%) | 11 ( 5.8%) |

Note:
Comparaisons at day 3,7,10 and 14 are considered for secondary analysis. The other days are used for exploratory analysis.


CONFIDENTIALITY STATEMENT: Information contained in this statistical report is confidential and should not be disclosed to other parties than those directly involved with the execution of this study without written authorization from Montreal Heart Institute

2 - Efficacy Analysis
2.2 - Secondary Analysis
Table 2.2.3c - Summary of and Statistical Analysis for Presence of Feverish or Chills at Day 1 to 14, ITT population

| | | Placebo N=109 | Hesperidin N=107 | All N=216 |
|---|---|---|---|---|
| DAY 10 | n | 100 | 90 | 190 |
| | No | 94 ( 94.0%) | 86 ( 95.6%) | 180 ( 94.7%) |
| | Yes | 6 ( 6.0%) | 4 ( 4.4%) | 10 ( 5.3%) |
| DAY 11 | n | 98 | 86 | 184 |
| | No | 93 ( 94.9%) | 83 ( 96.5%) | 176 ( 95.7%) |
| | Yes | 5 ( 5.1%) | 3 ( 3.5%) | 8 ( 4.3%) |
| DAY 12 | n | 99 | 85 | 184 |
| | No | 95 ( 96.0%) | 83 ( 97.6%) | 178 ( 96.7%) |
| | Yes | 4 ( 4.0%) | 2 ( 2.4%) | 6 ( 3.3%) |
| DAY 13 | n | 98 | 83 | 181 |
| | No | 96 ( 98.0%) | 82 ( 98.8%) | 178 ( 98.3%) |
| | Yes | 2 ( 2.0%) | 1 ( 1.2%) | 3 ( 1.7%) |
| DAY 14 | n | 95 | 79 | 174 |
| | No | 92 ( 96.8%) | 78 ( 98.7%) | 170 ( 97.7%) |
| | Yes | 3 ( 3.2%) | 1 ( 1.3%) | 4 ( 2.3%) |

Note:
Comparaisons at day 3,7,10 and 14 are considered for secondary analysis. The other days are used for exploratory analysis.


CONFIDENTIALITY STATEMENT: Information contained in this statistical report is confidential and should not be disclosed to other parties than those directly involved with the execution of this study without written authorization from Montreal Heart Institute

2 – Efficacy Analysis
2.2 – Secondary Analysis
Table 2.2.3c - Summary of and Statistical Analysis for Presence of Feverish or Chills at Day 1 to 14, ITT Population (continued)

Generalized linear mixed model (repeated binary logistic regression) with terms for treatment group, time and treatment group x time interaction

| Contrast | OR (95% CI) | P-value |
|---|---|---|
| Hesperidin vs Placebo at Day 3 | 1.20 (0.62; 2.34) | 0.5894 |
| Hesperidin vs Placebo at Day 7 | 1.14 (0.45; 2.89) | 0.7887 |
| Hesperidin vs Placebo at Day 10 | 0.73 (0.20; 2.70) | 0.6348 |
| Hesperidin vs Placebo at Day 14 | 0.39 (0.04; 3.92) | 0.4257 |


CONFIDENTIALITY STATEMENT: Information contained in this statistical report is confidential and should not be disclosed to other parties than those directly involved with the execution of this study without written authorization from Montreal Heart Institute

2 - Efficacy Analysis
2.2 - Secondary Analysis
Table 2.2.3d - Summary of and Statistical Analysis for Presence of Sore Throat at Day 1 to 14, ITT population

| | | Placebo N=109 | Hesperidin N=107 | All N=216 |
|---|---|---|---|---|
| DAY 1 | n | 107 | 105 | 212 |
| | No | 66 ( 61.7%) | 62 ( 59.0%) | 128 ( 60.4%) |
| | Yes | 41 ( 38.3%) | 43 ( 41.0%) | 84 ( 39.6%) |
| DAY 2 | n | 106 | 105 | 211 |
| | No | 71 ( 67.0%) | 65 ( 61.9%) | 136 ( 64.5%) |
| | Yes | 35 ( 33.0%) | 40 ( 38.1%) | 75 ( 35.5%) |
| DAY 3 | n | 103 | 102 | 205 |
| | No | 80 ( 77.7%) | 69 ( 67.6%) | 149 ( 72.7%) |
| | Yes | 23 ( 22.3%) | 33 ( 32.4%) | 56 ( 27.3%) |
| DAY 4 | n | 103 | 96 | 199 |
| | No | 80 ( 77.7%) | 69 ( 71.9%) | 149 ( 74.9%) |
| | Yes | 23 ( 22.3%) | 27 ( 28.1%) | 50 ( 25.1%) |
| DAY 5 | n | 102 | 95 | 197 |
| | No | 79 ( 77.5%) | 70 ( 73.7%) | 149 ( 75.6%) |
| | Yes | 23 ( 22.5%) | 25 ( 26.3%) | 48 ( 24.4%) |
| DAY 6 | n | 103 | 92 | 195 |
| | No | 86 ( 83.5%) | 76 ( 82.6%) | 162 ( 83.1%) |
| | Yes | 17 ( 16.5%) | 16 ( 17.4%) | 33 ( 16.9%) |
| DAY 7 | n | 102 | 91 | 193 |
| | No | 89 ( 87.3%) | 74 ( 81.3%) | 163 ( 84.5%) |
| | Yes | 13 ( 12.7%) | 17 ( 18.7%) | 30 ( 15.5%) |
| DAY 8 | n | 101 | 90 | 191 |
| | No | 88 ( 87.1%) | 78 ( 86.7%) | 166 ( 86.9%) |
| | Yes | 13 ( 12.9%) | 12 ( 13.3%) | 25 ( 13.1%) |
| DAY 9 | n | 100 | 89 | 189 |
| | No | 87 ( 87.0%) | 84 ( 94.4%) | 171 ( 90.5%) |
| | Yes | 13 ( 13.0%) | 5 ( 5.6%) | 18 ( 9.5%) |

Note:
Comparaisons at day 3,7,10 and 14 are considered for secondary analysis. The other days are used for exploratory analysis.


CONFIDENTIALITY STATEMENT: Information contained in this statistical report is confidential and should not be disclosed to other parties than those directly involved with the execution of this study without written authorization from Montreal Heart Institute

2 - Efficacy Analysis
2.2 - Secondary Analysis
Table 2.2.3d - Summary of and Statistical Analysis for Presence of Sore Throat at Day 1 to 14, ITT population

| | | Placebo N=109 | Hesperidin N=107 | All N=216 |
|---|---|---|---|---|
| DAY 10 | n | 100 | 90 | 190 |
| | No | 92 ( 92.0%) | 84 ( 93.3%) | 176 ( 92.6%) |
| | Yes | 8 ( 8.0%) | 6 ( 6.7%) | 14 ( 7.4%) |
| DAY 11 | n | 98 | 86 | 184 |
| | No | 90 ( 91.8%) | 82 ( 95.3%) | 172 ( 93.5%) |
| | Yes | 8 ( 8.2%) | 4 ( 4.7%) | 12 ( 6.5%) |
| DAY 12 | n | 99 | 85 | 184 |
| | No | 93 ( 93.9%) | 81 ( 95.3%) | 174 ( 94.6%) |
| | Yes | 6 ( 6.1%) | 4 ( 4.7%) | 10 ( 5.4%) |
| DAY 13 | n | 98 | 83 | 181 |
| | No | 93 ( 94.9%) | 81 ( 97.6%) | 174 ( 96.1%) |
| | Yes | 5 ( 5.1%) | 2 ( 2.4%) | 7 ( 3.9%) |
| DAY 14 | n | 95 | 79 | 174 |
| | No | 91 ( 95.8%) | 76 ( 96.2%) | 167 ( 96.0%) |
| | Yes | 4 ( 4.2%) | 3 ( 3.8%) | 7 ( 4.0%) |

Note:
Comparaisons at day 3,7,10 and 14 are considered for secondary analysis. The other days are used for exploratory analysis.


CONFIDENTIALITY STATEMENT: Information contained in this statistical report is confidential and should not be disclosed to other parties than those directly involved with the execution of this study without written authorization from Montreal Heart Institute

2 – Efficacy Analysis
2.2 – Secondary Analysis
Table 2.2.3d - Summary of and Statistical Analysis for Presence of Sore Throat at Day 1 to 14, ITT Population (continued)

Generalized linear mixed model (repeated binary logistic regression) with terms for treatment group, time and treatment group x time interaction

| Contrast | OR (95% CI) | P-value |
|---|---|---|
| Hesperidin vs Placebo at Day 3 | 1.66 (0.89; 3.11) | 0.1113 |
| Hesperidin vs Placebo at Day 7 | 1.57 (0.71; 3.47) | 0.2613 |
| Hesperidin vs Placebo at Day 10 | 0.82 (0.27; 2.49) | 0.7273 |
| Hesperidin vs Placebo at Day 14 | 0.90 (0.19; 4.19) | 0.8909 |


CONFIDENTIALITY STATEMENT: Information contained in this statistical report is confidential and should not be disclosed to other parties than those directly involved with the execution of this study without written authorization from Montreal Heart Institute

2 – Efficacy Analysis
2.2 – Secondary Analysis
Table 2.2.3e – Summary of and Statistical Analysis for Presence of Runny Nose at Day 1 to 14, ITT population

| | | Placebo N=109 | Hesperidin N=107 | All N=216 |
|---|---|---|---|---|
| DAY 1 | n | 107 | 105 | 212 |
| | No | 57 ( 53.3%) | 52 ( 49.5%) | 109 ( 51.4%) |
| | Yes | 50 ( 46.7%) | 53 ( 50.5%) | 103 ( 48.6%) |
| DAY 2 | n | 106 | 105 | 211 |
| | No | 59 ( 55.7%) | 50 ( 47.6%) | 109 ( 51.7%) |
| | Yes | 47 ( 44.3%) | 55 ( 52.4%) | 102 ( 48.3%) |
| DAY 3 | n | 103 | 102 | 205 |
| | No | 65 ( 63.1%) | 54 ( 52.9%) | 119 ( 58.0%) |
| | Yes | 38 ( 36.9%) | 48 ( 47.1%) | 86 ( 42.0%) |
| DAY 4 | n | 103 | 96 | 199 |
| | No | 68 ( 66.0%) | 56 ( 58.3%) | 124 ( 62.3%) |
| | Yes | 35 ( 34.0%) | 40 ( 41.7%) | 75 ( 37.7%) |
| DAY 5 | n | 102 | 95 | 197 |
| | No | 70 ( 68.6%) | 59 ( 62.1%) | 129 ( 65.5%) |
| | Yes | 32 ( 31.4%) | 36 ( 37.9%) | 68 ( 34.5%) |
| DAY 6 | n | 103 | 92 | 195 |
| | No | 74 ( 71.8%) | 64 ( 69.6%) | 138 ( 70.8%) |
| | Yes | 29 ( 28.2%) | 28 ( 30.4%) | 57 ( 29.2%) |
| DAY 7 | n | 102 | 91 | 193 |
| | No | 80 ( 78.4%) | 66 ( 72.5%) | 146 ( 75.6%) |
| | Yes | 22 ( 21.6%) | 25 ( 27.5%) | 47 ( 24.4%) |
| DAY 8 | n | 101 | 90 | 191 |
| | No | 78 ( 77.2%) | 66 ( 73.3%) | 144 ( 75.4%) |
| | Yes | 23 ( 22.8%) | 24 ( 26.7%) | 47 ( 24.6%) |
| DAY 9 | n | 100 | 89 | 189 |
| | No | 78 ( 78.0%) | 68 ( 76.4%) | 146 ( 77.2%) |
| | Yes | 22 ( 22.0%) | 21 ( 23.6%) | 43 ( 22.8%) |

Note:
Comparaisons at day 3,7,10 and 14 are considered for secondary analysis. The other days are used for exploratory analysis.


CONFIDENTIALITY STATEMENT: Information contained in this statistical report is confidential and should not be disclosed to other parties than those directly involved with the execution of this study without written authorization from Montreal Heart Institute

2 - Efficacy Analysis
2.2 - Secondary Analysis
Table 2.2.3e - Summary of and Statistical Analysis for Presence of Runny Nose at Day 1 to 14, ITT population

| | | Placebo N=109 | Hesperidin N=107 | All N=216 |
|---|---|---|---|---|
| DAY 10 | n | 100 | 90 | 190 |
| | No | 83 ( 83.0%) | 72 ( 80.0%) | 155 ( 81.6%) |
| | Yes | 17 ( 17.0%) | 18 ( 20.0%) | 35 ( 18.4%) |
| DAY 11 | n | 98 | 86 | 184 |
| | No | 83 ( 84.7%) | 71 ( 82.6%) | 154 ( 83.7%) |
| | Yes | 15 ( 15.3%) | 15 ( 17.4%) | 30 ( 16.3%) |
| DAY 12 | n | 99 | 85 | 184 |
| | No | 86 ( 86.9%) | 72 ( 84.7%) | 158 ( 85.9%) |
| | Yes | 13 ( 13.1%) | 13 ( 15.3%) | 26 ( 14.1%) |
| DAY 13 | n | 98 | 83 | 181 |
| | No | 86 ( 87.8%) | 69 ( 83.1%) | 155 ( 85.6%) |
| | Yes | 12 ( 12.2%) | 14 ( 16.9%) | 26 ( 14.4%) |
| DAY 14 | n | 95 | 79 | 174 |
| | No | 85 ( 89.5%) | 67 ( 84.8%) | 152 ( 87.4%) |
| | Yes | 10 ( 10.5%) | 12 ( 15.2%) | 22 ( 12.6%) |

Note:
Comparaisons at day 3,7,10 and 14 are considered for secondary analysis. The other days are used for exploratory analysis.


CONFIDENTIALITY STATEMENT: Information contained in this statistical report is confidential and should not be disclosed to other parties than those directly involved with the execution of this study without written authorization from Montreal Heart Institute

2 – Efficacy Analysis
2.2 – Secondary Analysis
Table 2.2.3e - Summary of and Statistical Analysis for Presence of Runny Nose at Day 1 to 14, ITT Population (continued)

Generalized linear mixed model (repeated binary logistic regression) with terms for treatment group, time and treatment group x time interaction

| Contrast | OR (95% CI) | P-value |
|---|---|---|
| Hesperidin vs Placebo at Day 3 | 1.52 (0.87; 2.67) | 0.1438 |
| Hesperidin vs Placebo at Day 7 | 1.38 (0.71; 2.68) | 0.3440 |
| Hesperidin vs Placebo at Day 10 | 1.22 (0.58; 2.56) | 0.5968 |
| Hesperidin vs Placebo at Day 14 | 1.52 (0.62; 3.76) | 0.3620 |


CONFIDENTIALITY STATEMENT: Information contained in this statistical report is confidential and should not be disclosed to other parties than those directly involved with the execution of this study without written authorization from Montreal Heart Institute

2 - Efficacy Analysis
2.2 - Secondary Analysis
Table 2.2.3f - Summary of and Statistical Analysis for Presence of Shortness of Breath/Difficulty Breathing at Day 1 to 14, ITT population

| | | Placebo N=109 | Hesperidin N=107 | All N=216 |
|---|---|---|---|---|
| DAY 1 | n | 107 | 105 | 212 |
| | No | 65 ( 60.7%) | 54 ( 51.4%) | 119 ( 56.1%) |
| | Yes | 42 ( 39.3%) | 51 ( 48.6%) | 93 ( 43.9%) |
| DAY 2 | n | 106 | 105 | 211 |
| | No | 62 ( 58.5%) | 52 ( 49.5%) | 114 ( 54.0%) |
| | Yes | 44 ( 41.5%) | 53 ( 50.5%) | 97 ( 46.0%) |
| DAY 3 | n | 103 | 102 | 205 |
| | No | 68 ( 66.0%) | 56 ( 54.9%) | 124 ( 60.5%) |
| | Yes | 35 ( 34.0%) | 46 ( 45.1%) | 81 ( 39.5%) |
| DAY 4 | n | 103 | 96 | 199 |
| | No | 68 ( 66.0%) | 58 ( 60.4%) | 126 ( 63.3%) |
| | Yes | 35 ( 34.0%) | 38 ( 39.6%) | 73 ( 36.7%) |
| DAY 5 | n | 102 | 95 | 197 |
| | No | 71 ( 69.6%) | 56 ( 58.9%) | 127 ( 64.5%) |
| | Yes | 31 ( 30.4%) | 39 ( 41.1%) | 70 ( 35.5%) |
| DAY 6 | n | 103 | 92 | 195 |
| | No | 76 ( 73.8%) | 56 ( 60.9%) | 132 ( 67.7%) |
| | Yes | 27 ( 26.2%) | 36 ( 39.1%) | 63 ( 32.3%) |
| DAY 7 | n | 102 | 91 | 193 |
| | No | 72 ( 70.6%) | 63 ( 69.2%) | 135 ( 69.9%) |
| | Yes | 30 ( 29.4%) | 28 ( 30.8%) | 58 ( 30.1%) |
| DAY 8 | n | 101 | 90 | 191 |
| | No | 73 ( 72.3%) | 63 ( 70.0%) | 136 ( 71.2%) |
| | Yes | 28 ( 27.7%) | 27 ( 30.0%) | 55 ( 28.8%) |
| DAY 9 | n | 100 | 89 | 189 |
| | No | 79 ( 79.0%) | 64 ( 71.9%) | 143 ( 75.7%) |
| | Yes | 21 ( 21.0%) | 25 ( 28.1%) | 46 ( 24.3%) |

Note:
Comparaisons at day 3,7,10 and 14 are considered for secondary analysis. The other days are used for exploratory analysis.


CONFIDENTIALITY STATEMENT: Information contained in this statistical report is confidential and should not be disclosed to other parties than those directly involved with the execution of this study without written authorization from Montreal Heart Institute

2 - Efficacy Analysis
2.2 - Secondary Analysis
Table 2.2.3f - Summary of and Statistical Analysis for Presence of Shortness of Breath/Difficulty Breathing at Day 1 to 14, ITT population

| | | Placebo N=109 | Hesperidin N=107 | All N=216 |
|---|---|---|---|---|
| DAY 10 | n | 100 | 90 | 190 |
| | No | 82 ( 82.0%) | 71 ( 78.9%) | 153 ( 80.5%) |
| | Yes | 18 ( 18.0%) | 19 ( 21.1%) | 37 ( 19.5%) |
| DAY 11 | n | 98 | 86 | 184 |
| | No | 80 ( 81.6%) | 70 ( 81.4%) | 150 ( 81.5%) |
| | Yes | 18 ( 18.4%) | 16 ( 18.6%) | 34 ( 18.5%) |
| DAY 12 | n | 99 | 85 | 184 |
| | No | 84 ( 84.8%) | 73 ( 85.9%) | 157 ( 85.3%) |
| | Yes | 15 ( 15.2%) | 12 ( 14.1%) | 27 ( 14.7%) |
| DAY 13 | n | 98 | 83 | 181 |
| | No | 83 ( 84.7%) | 70 ( 84.3%) | 153 ( 84.5%) |
| | Yes | 15 ( 15.3%) | 13 ( 15.7%) | 28 ( 15.5%) |
| DAY 14 | n | 95 | 79 | 174 |
| | No | 82 ( 86.3%) | 65 ( 82.3%) | 147 ( 84.5%) |
| | Yes | 13 ( 13.7%) | 14 ( 17.7%) | 27 ( 15.5%) |

Note:
Comparaisons at day 3,7,10 and 14 are considered for secondary analysis. The other days are used for exploratory analysis.


CONFIDENTIALITY STATEMENT: Information contained in this statistical report is confidential and should not be disclosed to other parties than those directly involved with the execution of this study without written authorization from Montreal Heart Institute

2 – Efficacy Analysis
2.2 – Secondary Analysis
Table 2.2.3f - Summary of and Statistical Analysis for Presence of Shortness of Breath/Difficulty Breathing at Day 1 to 14, ITT Population (continued)

Generalized linear mixed model (repeated binary logistic regression) with terms for treatment group, time and treatment group x time interaction

| Contrast | OR (95% CI) | P-value |
|---|---|---|
| Hesperidin vs Placebo at Day 3 | 1.60 (0.90; 2.82) | 0.1068 |
| Hesperidin vs Placebo at Day 7 | 1.07 (0.57; 1.98) | 0.8382 |
| Hesperidin vs Placebo at Day 10 | 1.22 (0.59; 2.51) | 0.5912 |
| Hesperidin vs Placebo at Day 14 | 1.36 (0.59; 3.11) | 0.4677 |


CONFIDENTIALITY STATEMENT: Information contained in this statistical report is confidential and should not be disclosed to other parties than those directly involved with the execution of this study without written authorization from Montreal Heart Institute

2 - Efficacy Analysis
2.2 - Secondary Analysis
Table 2.2.3g - Summary of and Statistical Analysis for Presence of Nausea/Vomiting at Day 1 to 14, ITT population

| | | Placebo N=109 | Hesperidin N=107 | All N=216 |
|---|---|---|---|---|
| DAY 1 | n | 107 | 105 | 212 |
| | No | 95 ( 88.8%) | 83 ( 79.0%) | 178 ( 84.0%) |
| | Yes | 12 ( 11.2%) | 22 ( 21.0%) | 34 ( 16.0%) |
| DAY 2 | n | 106 | 105 | 211 |
| | No | 91 ( 85.8%) | 83 ( 79.0%) | 174 ( 82.5%) |
| | Yes | 15 ( 14.2%) | 22 ( 21.0%) | 37 ( 17.5%) |
| DAY 3 | n | 103 | 102 | 205 |
| | No | 91 ( 88.3%) | 81 ( 79.4%) | 172 ( 83.9%) |
| | Yes | 12 ( 11.7%) | 21 ( 20.6%) | 33 ( 16.1%) |
| DAY 4 | n | 103 | 96 | 199 |
| | No | 94 ( 91.3%) | 81 ( 84.4%) | 175 ( 87.9%) |
| | Yes | 9 ( 8.7%) | 15 ( 15.6%) | 24 ( 12.1%) |
| DAY 5 | n | 102 | 95 | 197 |
| | No | 95 ( 93.1%) | 80 ( 84.2%) | 175 ( 88.8%) |
| | Yes | 7 ( 6.9%) | 15 ( 15.8%) | 22 ( 11.2%) |
| DAY 6 | n | 103 | 92 | 195 |
| | No | 97 ( 94.2%) | 79 ( 85.9%) | 176 ( 90.3%) |
| | Yes | 6 ( 5.8%) | 13 ( 14.1%) | 19 ( 9.7%) |
| DAY 7 | n | 102 | 91 | 193 |
| | No | 91 ( 89.2%) | 82 ( 90.1%) | 173 ( 89.6%) |
| | Yes | 11 ( 10.8%) | 9 ( 9.9%) | 20 ( 10.4%) |
| DAY 8 | n | 101 | 90 | 191 |
| | No | 95 ( 94.1%) | 81 ( 90.0%) | 176 ( 92.1%) |
| | Yes | 6 ( 5.9%) | 9 ( 10.0%) | 15 ( 7.9%) |
| DAY 9 | n | 100 | 89 | 189 |
| | No | 96 ( 96.0%) | 81 ( 91.0%) | 177 ( 93.7%) |
| | Yes | 4 ( 4.0%) | 8 ( 9.0%) | 12 ( 6.3%) |

Note:
Comparaisons at day 3,7,10 and 14 are considered for secondary analysis. The other days are used for exploratory analysis.


CONFIDENTIALITY STATEMENT: Information contained in this statistical report is confidential and should not be disclosed to other parties than those directly involved with the execution of this study without written authorization from Montreal Heart Institute

2 - Efficacy Analysis
2.2 - Secondary Analysis
Table 2.2.3g - Summary of and Statistical Analysis for Presence of Nausea/Vomiting at Day 1 to 14, ITT population

| | | Placebo N=109 | Hesperidin N=107 | All N=216 |
|---|---|---|---|---|
| DAY 10 | n | 100 | 90 | 190 |
| | No | 95 ( 95.0%) | 84 ( 93.3%) | 179 ( 94.2%) |
| | Yes | 5 ( 5.0%) | 6 ( 6.7%) | 11 ( 5.8%) |
| DAY 11 | n | 98 | 86 | 184 |
| | No | 93 ( 94.9%) | 82 ( 95.3%) | 175 ( 95.1%) |
| | Yes | 5 ( 5.1%) | 4 ( 4.7%) | 9 ( 4.9%) |
| DAY 12 | n | 99 | 85 | 184 |
| | No | 96 ( 97.0%) | 83 ( 97.6%) | 179 ( 97.3%) |
| | Yes | 3 ( 3.0%) | 2 ( 2.4%) | 5 ( 2.7%) |
| DAY 13 | n | 98 | 83 | 181 |
| | No | 94 ( 95.9%) | 82 ( 98.8%) | 176 ( 97.2%) |
| | Yes | 4 ( 4.1%) | 1 ( 1.2%) | 5 ( 2.8%) |
| DAY 14 | n | 95 | 79 | 174 |
| | No | 93 ( 97.9%) | 77 ( 97.5%) | 170 ( 97.7%) |
| | Yes | 2 ( 2.1%) | 2 ( 2.5%) | 4 ( 2.3%) |

Note:
Comparaisons at day 3,7,10 and 14 are considered for secondary analysis. The other days are used for exploratory analysis.


CONFIDENTIALITY STATEMENT: Information contained in this statistical report is confidential and should not be disclosed to other parties than those directly involved with the execution of this study without written authorization from Montreal Heart Institute

2 – Efficacy Analysis
2.2 – Secondary Analysis
Table 2.2.3g - Summary of and Statistical Analysis for Presence of Nausea/Vomiting at Day 1 to 14, ITT Population (continued)

Generalized linear mixed model (repeated binary logistic regression) with terms for treatment group, time and treatment group x time interaction

| Contrast | OR (95% CI) | P-value |
|---|---|---|
| Hesperidin vs Placebo at Day 3 | 1.97 (0.91; 4.27) | 0.0875 |
| Hesperidin vs Placebo at Day 7 | 0.91 (0.36; 2.32) | 0.8397 |
| Hesperidin vs Placebo at Day 10 | 1.36 (0.40; 4.65) | 0.6265 |
| Hesperidin vs Placebo at Day 14 | 1.21 (0.16; 8.91) | 0.8528 |


CONFIDENTIALITY STATEMENT: Information contained in this statistical report is confidential and should not be disclosed to other parties than those directly involved with the execution of this study without written authorization from Montreal Heart Institute

2 - Efficacy Analysis
2.2 - Secondary Analysis
Table 2.2.3h - Summary of and Statistical Analysis for Presence of Headache at Day 1 to 14, ITT population

| | | Placebo N=109 | Hesperidin N=107 | All N=216 |
|---|---|---|---|---|
| DAY 1 | n | 107 | 105 | 212 |
| | No | 39 ( 36.4%) | 38 ( 36.2%) | 77 ( 36.3%) |
| | Yes | 68 ( 63.6%) | 67 ( 63.8%) | 135 ( 63.7%) |
| DAY 2 | n | 106 | 105 | 211 |
| | No | 54 ( 50.9%) | 49 ( 46.7%) | 103 ( 48.8%) |
| | Yes | 52 ( 49.1%) | 56 ( 53.3%) | 108 ( 51.2%) |
| DAY 3 | n | 103 | 102 | 205 |
| | No | 62 ( 60.2%) | 54 ( 52.9%) | 116 ( 56.6%) |
| | Yes | 41 ( 39.8%) | 48 ( 47.1%) | 89 ( 43.4%) |
| DAY 4 | n | 103 | 96 | 199 |
| | No | 68 ( 66.0%) | 53 ( 55.2%) | 121 ( 60.8%) |
| | Yes | 35 ( 34.0%) | 43 ( 44.8%) | 78 ( 39.2%) |
| DAY 5 | n | 102 | 95 | 197 |
| | No | 66 ( 64.7%) | 58 ( 61.1%) | 124 ( 62.9%) |
| | Yes | 36 ( 35.3%) | 37 ( 38.9%) | 73 ( 37.1%) |
| DAY 6 | n | 103 | 92 | 195 |
| | No | 73 ( 70.9%) | 58 ( 63.0%) | 131 ( 67.2%) |
| | Yes | 30 ( 29.1%) | 34 ( 37.0%) | 64 ( 32.8%) |
| DAY 7 | n | 102 | 91 | 193 |
| | No | 68 ( 66.7%) | 57 ( 62.6%) | 125 ( 64.8%) |
| | Yes | 34 ( 33.3%) | 34 ( 37.4%) | 68 ( 35.2%) |
| DAY 8 | n | 101 | 90 | 191 |
| | No | 74 ( 73.3%) | 68 ( 75.6%) | 142 ( 74.3%) |
| | Yes | 27 ( 26.7%) | 22 ( 24.4%) | 49 ( 25.7%) |
| DAY 9 | n | 100 | 89 | 189 |
| | No | 79 ( 79.0%) | 70 ( 78.7%) | 149 ( 78.8%) |
| | Yes | 21 ( 21.0%) | 19 ( 21.3%) | 40 ( 21.2%) |

Note:
Comparaisons at day 3,7,10 and 14 are considered for secondary analysis. The other days are used for exploratory analysis.


CONFIDENTIALITY STATEMENT: Information contained in this statistical report is confidential and should not be disclosed to other parties than those directly involved with the execution of this study without written authorization from Montreal Heart Institute

2 - Efficacy Analysis
2.2 - Secondary Analysis
Table 2.2.3h - Summary of and Statistical Analysis for Presence of Headache at Day 1 to 14, ITT population

| | | Placebo N=109 | Hesperidin N=107 | All N=216 |
|---|---|---|---|---|
| DAY 10 | n | 100 | 90 | 190 |
| | No | 80 ( 80.0%) | 68 ( 75.6%) | 148 ( 77.9%) |
| | Yes | 20 ( 20.0%) | 22 ( 24.4%) | 42 ( 22.1%) |
| DAY 11 | n | 98 | 86 | 184 |
| | No | 81 ( 82.7%) | 70 ( 81.4%) | 151 ( 82.1%) |
| | Yes | 17 ( 17.3%) | 16 ( 18.6%) | 33 ( 17.9%) |
| DAY 12 | n | 99 | 85 | 184 |
| | No | 82 ( 82.8%) | 73 ( 85.9%) | 155 ( 84.2%) |
| | Yes | 17 ( 17.2%) | 12 ( 14.1%) | 29 ( 15.8%) |
| DAY 13 | n | 98 | 83 | 181 |
| | No | 84 ( 85.7%) | 74 ( 89.2%) | 158 ( 87.3%) |
| | Yes | 14 ( 14.3%) | 9 ( 10.8%) | 23 ( 12.7%) |
| DAY 14 | n | 95 | 79 | 174 |
| | No | 86 ( 90.5%) | 69 ( 87.3%) | 155 ( 89.1%) |
| | Yes | 9 ( 9.5%) | 10 ( 12.7%) | 19 ( 10.9%) |

Note:
Comparaisons at day 3,7,10 and 14 are considered for secondary analysis. The other days are used for exploratory analysis.


CONFIDENTIALITY STATEMENT: Information contained in this statistical report is confidential and should not be disclosed to other parties than those directly involved with the execution of this study without written authorization from Montreal Heart Institute

2 – Efficacy Analysis
2.2 – Secondary Analysis
Table 2.2.3h - Summary of and Statistical Analysis for Presence of Headache at Day 1 to 14, ITT Population (continued)

Generalized linear mixed model (repeated binary logistic regression) with terms for treatment group, time and treatment group x time interaction

| Contrast | OR (95% CI) | P-value |
|---|---|---|
| Hesperidin vs Placebo at Day 3 | 1.34 (0.77; 2.35) | 0.2983 |
| Hesperidin vs Placebo at Day 7 | 1.19 (0.66; 2.16) | 0.5611 |
| Hesperidin vs Placebo at Day 10 | 1.29 (0.65; 2.58) | 0.4643 |
| Hesperidin vs Placebo at Day 14 | 1.38 (0.53; 3.62) | 0.5063 |


CONFIDENTIALITY STATEMENT: Information contained in this statistical report is confidential and should not be disclosed to other parties than those directly involved with the execution of this study without written authorization from Montreal Heart Institute

2 - Efficacy Analysis
2.2 - Secondary Analysis
Table 2.2.3i - Summary of and Statistical Analysis for Presence of General Weakness at Day 1 to 14, ITT population

| | | Placebo N=109 | Hesperidin N=107 | All N=216 |
|---|---|---|---|---|
| DAY 1 | n | 107 | 105 | 212 |
| | No | 40 ( 37.4%) | 32 ( 30.5%) | 72 ( 34.0%) |
| | Yes | 67 ( 62.6%) | 73 ( 69.5%) | 140 ( 66.0%) |
| DAY 2 | n | 106 | 105 | 211 |
| | No | 45 ( 42.5%) | 35 ( 33.3%) | 80 ( 37.9%) |
| | Yes | 61 ( 57.5%) | 70 ( 66.7%) | 131 ( 62.1%) |
| DAY 3 | n | 103 | 102 | 205 |
| | No | 48 ( 46.6%) | 39 ( 38.2%) | 87 ( 42.4%) |
| | Yes | 55 ( 53.4%) | 63 ( 61.8%) | 118 ( 57.6%) |
| DAY 4 | n | 103 | 96 | 199 |
| | No | 49 ( 47.6%) | 37 ( 38.5%) | 86 ( 43.2%) |
| | Yes | 54 ( 52.4%) | 59 ( 61.5%) | 113 ( 56.8%) |
| DAY 5 | n | 102 | 95 | 197 |
| | No | 53 ( 52.0%) | 41 ( 43.2%) | 94 ( 47.7%) |
| | Yes | 49 ( 48.0%) | 54 ( 56.8%) | 103 ( 52.3%) |
| DAY 6 | n | 103 | 92 | 195 |
| | No | 62 ( 60.2%) | 47 ( 51.1%) | 109 ( 55.9%) |
| | Yes | 41 ( 39.8%) | 45 ( 48.9%) | 86 ( 44.1%) |
| DAY 7 | n | 102 | 91 | 193 |
| | No | 62 ( 60.8%) | 52 ( 57.1%) | 114 ( 59.1%) |
| | Yes | 40 ( 39.2%) | 39 ( 42.9%) | 79 ( 40.9%) |
| DAY 8 | n | 101 | 90 | 191 |
| | No | 69 ( 68.3%) | 56 ( 62.2%) | 125 ( 65.4%) |
| | Yes | 32 ( 31.7%) | 34 ( 37.8%) | 66 ( 34.6%) |
| DAY 9 | n | 100 | 89 | 189 |
| | No | 73 ( 73.0%) | 62 ( 69.7%) | 135 ( 71.4%) |
| | Yes | 27 ( 27.0%) | 27 ( 30.3%) | 54 ( 28.6%) |

Note:
Comparaisons at day 3,7,10 and 14 are considered for secondary analysis. The other days are used for exploratory analysis.


CONFIDENTIALITY STATEMENT: Information contained in this statistical report is confidential and should not be disclosed to other parties than those directly involved with the execution of this study without written authorization from Montreal Heart Institute

2 - Efficacy Analysis
2.2 - Secondary Analysis
Table 2.2.3i - Summary of and Statistical Analysis for Presence of General Weakness at Day 1 to 14, ITT population

| | | Placebo N=109 | Hesperidin N=107 | All N=216 |
|---|---|---|---|---|
| DAY 10 | n | 100 | 90 | 190 |
| | No | 79 ( 79.0%) | 70 ( 77.8%) | 149 ( 78.4%) |
| | Yes | 21 ( 21.0%) | 20 ( 22.2%) | 41 ( 21.6%) |
| DAY 11 | n | 98 | 86 | 184 |
| | No | 78 ( 79.6%) | 67 ( 77.9%) | 145 ( 78.8%) |
| | Yes | 20 ( 20.4%) | 19 ( 22.1%) | 39 ( 21.2%) |
| DAY 12 | n | 99 | 85 | 184 |
| | No | 78 ( 78.8%) | 69 ( 81.2%) | 147 ( 79.9%) |
| | Yes | 21 ( 21.2%) | 16 ( 18.8%) | 37 ( 20.1%) |
| DAY 13 | n | 98 | 83 | 181 |
| | No | 78 ( 79.6%) | 68 ( 81.9%) | 146 ( 80.7%) |
| | Yes | 20 ( 20.4%) | 15 ( 18.1%) | 35 ( 19.3%) |
| DAY 14 | n | 95 | 79 | 174 |
| | No | 78 ( 82.1%) | 65 ( 82.3%) | 143 ( 82.2%) |
| | Yes | 17 ( 17.9%) | 14 ( 17.7%) | 31 ( 17.8%) |

Note:
Comparaisons at day 3,7,10 and 14 are considered for secondary analysis. The other days are used for exploratory analysis.


CONFIDENTIALITY STATEMENT: Information contained in this statistical report is confidential and should not be disclosed to other parties than those directly involved with the execution of this study without written authorization from Montreal Heart Institute

2 – Efficacy Analysis
2.2 – Secondary Analysis
Table 2.2.3i - Summary of and Statistical Analysis for Presence of General Weakness at Day 1 to 14, ITT Population (continued)

Generalized linear mixed model (repeated binary logistic regression) with terms for treatment group, time and treatment group x time interaction

| Contrast | OR (95% CI) | P-value |
|---|---|---|
| Hesperidin vs Placebo at Day 3 | 1.41 (0.80; 2.47) | 0.2292 |
| Hesperidin vs Placebo at Day 7 | 1.16 (0.65; 2.07) | 0.6097 |
| Hesperidin vs Placebo at Day 10 | 1.07 (0.54; 2.16) | 0.8389 |
| Hesperidin vs Placebo at Day 14 | 0.99 (0.45; 2.17) | 0.9764 |


CONFIDENTIALITY STATEMENT: Information contained in this statistical report is confidential and should not be disclosed to other parties than those directly involved with the execution of this study without written authorization from Montreal Heart Institute

2 – Efficacy Analysis
2.2 – Secondary Analysis
Table 2.2.3j – Summary of and Statistical Analysis for Presence of Pain at Day 1 to 14, ITT population

| | | Placebo N=109 | Hesperidin N=107 | All N=216 |
|---|---|---|---|---|
| DAY 1 | n | 107 | 105 | 212 |
| | No | 44 ( 41.1%) | 43 ( 41.0%) | 87 ( 41.0%) |
| | Yes | 63 ( 58.9%) | 62 ( 59.0%) | 125 ( 59.0%) |
| DAY 2 | n | 106 | 105 | 211 |
| | No | 55 ( 51.9%) | 47 ( 44.8%) | 102 ( 48.3%) |
| | Yes | 51 ( 48.1%) | 58 ( 55.2%) | 109 ( 51.7%) |
| DAY 3 | n | 103 | 102 | 205 |
| | No | 53 ( 51.5%) | 58 ( 56.9%) | 111 ( 54.1%) |
| | Yes | 50 ( 48.5%) | 44 ( 43.1%) | 94 ( 45.9%) |
| DAY 4 | n | 103 | 96 | 199 |
| | No | 65 ( 63.1%) | 63 ( 65.6%) | 128 ( 64.3%) |
| | Yes | 38 ( 36.9%) | 33 ( 34.4%) | 71 ( 35.7%) |
| DAY 5 | n | 102 | 95 | 197 |
| | No | 69 ( 67.6%) | 68 ( 71.6%) | 137 ( 69.5%) |
| | Yes | 33 ( 32.4%) | 27 ( 28.4%) | 60 ( 30.5%) |
| DAY 6 | n | 103 | 92 | 195 |
| | No | 79 ( 76.7%) | 71 ( 77.2%) | 150 ( 76.9%) |
| | Yes | 24 ( 23.3%) | 21 ( 22.8%) | 45 ( 23.1%) |
| DAY 7 | n | 102 | 91 | 193 |
| | No | 79 ( 77.5%) | 74 ( 81.3%) | 153 ( 79.3%) |
| | Yes | 23 ( 22.5%) | 17 ( 18.7%) | 40 ( 20.7%) |
| DAY 8 | n | 101 | 90 | 191 |
| | No | 82 ( 81.2%) | 77 ( 85.6%) | 159 ( 83.2%) |
| | Yes | 19 ( 18.8%) | 13 ( 14.4%) | 32 ( 16.8%) |
| DAY 9 | n | 100 | 89 | 189 |
| | No | 84 ( 84.0%) | 76 ( 85.4%) | 160 ( 84.7%) |
| | Yes | 16 ( 16.0%) | 13 ( 14.6%) | 29 ( 15.3%) |

Note:
Comparaisons at day 3,7,10 and 14 are considered for secondary analysis. The other days are used for exploratory analysis.


CONFIDENTIALITY STATEMENT: Information contained in this statistical report is confidential and should not be disclosed to other parties than those directly involved with the execution of this study without written authorization from Montreal Heart Institute

2 - Efficacy Analysis
2.2 - Secondary Analysis
Table 2.2.3j - Summary of and Statistical Analysis for Presence of Pain at Day 1 to 14, ITT population

| | | Placebo N=109 | Hesperidin N=107 | All N=216 |
|---|---|---|---|---|
| DAY 10 | n | 100 | 90 | 190 |
| | No | 86 ( 86.0%) | 77 ( 85.6%) | 163 ( 85.8%) |
| | Yes | 14 ( 14.0%) | 13 ( 14.4%) | 27 ( 14.2%) |
| DAY 11 | n | 98 | 86 | 184 |
| | No | 86 ( 87.8%) | 77 ( 89.5%) | 163 ( 88.6%) |
| | Yes | 12 ( 12.2%) | 9 ( 10.5%) | 21 ( 11.4%) |
| DAY 12 | n | 99 | 85 | 184 |
| | No | 89 ( 89.9%) | 79 ( 92.9%) | 168 ( 91.3%) |
| | Yes | 10 ( 10.1%) | 6 ( 7.1%) | 16 ( 8.7%) |
| DAY 13 | n | 98 | 83 | 181 |
| | No | 89 ( 90.8%) | 78 ( 94.0%) | 167 ( 92.3%) |
| | Yes | 9 ( 9.2%) | 5 ( 6.0%) | 14 ( 7.7%) |
| DAY 14 | n | 95 | 79 | 174 |
| | No | 87 ( 91.6%) | 74 ( 93.7%) | 161 ( 92.5%) |
| | Yes | 8 ( 8.4%) | 5 ( 6.3%) | 13 ( 7.5%) |

Note:
Comparaisons at day 3,7,10 and 14 are considered for secondary analysis. The other days are used for exploratory analysis.


CONFIDENTIALITY STATEMENT: Information contained in this statistical report is confidential and should not be disclosed to other parties than those directly involved with the execution of this study without written authorization from Montreal Heart Institute

2 – Efficacy Analysis
2.2 – Secondary Analysis
Table 2.2.3j - Summary of and Statistical Analysis for Presence of Pain at Day 1 to 14, ITT Population (continued)

Generalized linear mixed model (repeated binary logistic regression) with terms for treatment group, time and treatment group x time interaction

| Contrast | OR (95% CI) | P-value |
|---|---|---|
| Hesperidin vs Placebo at Day 3 | 0.80 (0.46; 1.40) | 0.4403 |
| Hesperidin vs Placebo at Day 7 | 0.79 (0.39; 1.60) | 0.5112 |
| Hesperidin vs Placebo at Day 10 | 1.04 (0.46; 2.36) | 0.9306 |
| Hesperidin vs Placebo at Day 14 | 0.73 (0.23; 2.36) | 0.6046 |


CONFIDENTIALITY STATEMENT: Information contained in this statistical report is confidential and should not be disclosed to other parties than those directly involved with the execution of this study without written authorization from Montreal Heart Institute

2 - Efficacy Analysis
2.2 - Secondary Analysis
Table 2.2.3k - Summary of and Statistical Analysis for Presence of Irritability/Confusion at Day 1 to 14, ITT population

| | | Placebo N=109 | Hesperidin N=107 | All N=216 |
|---|---|---|---|---|
| DAY 1 | n | 107 | 105 | 212 |
| | No | 79 ( 73.8%) | 80 ( 76.2%) | 159 ( 75.0%) |
| | Yes | 28 ( 26.2%) | 25 ( 23.8%) | 53 ( 25.0%) |
| DAY 2 | n | 106 | 105 | 211 |
| | No | 80 ( 75.5%) | 83 ( 79.0%) | 163 ( 77.3%) |
| | Yes | 26 ( 24.5%) | 22 ( 21.0%) | 48 ( 22.7%) |
| DAY 3 | n | 103 | 102 | 205 |
| | No | 85 ( 82.5%) | 82 ( 80.4%) | 167 ( 81.5%) |
| | Yes | 18 ( 17.5%) | 20 ( 19.6%) | 38 ( 18.5%) |
| DAY 4 | n | 103 | 96 | 199 |
| | No | 83 ( 80.6%) | 77 ( 80.2%) | 160 ( 80.4%) |
| | Yes | 20 ( 19.4%) | 19 ( 19.8%) | 39 ( 19.6%) |
| DAY 5 | n | 102 | 95 | 197 |
| | No | 87 ( 85.3%) | 85 ( 89.5%) | 172 ( 87.3%) |
| | Yes | 15 ( 14.7%) | 10 ( 10.5%) | 25 ( 12.7%) |
| DAY 6 | n | 103 | 92 | 195 |
| | No | 91 ( 88.3%) | 81 ( 88.0%) | 172 ( 88.2%) |
| | Yes | 12 ( 11.7%) | 11 ( 12.0%) | 23 ( 11.8%) |
| DAY 7 | n | 102 | 91 | 193 |
| | No | 92 ( 90.2%) | 85 ( 93.4%) | 177 ( 91.7%) |
| | Yes | 10 ( 9.8%) | 6 ( 6.6%) | 16 ( 8.3%) |
| DAY 8 | n | 101 | 90 | 191 |
| | No | 92 ( 91.1%) | 85 ( 94.4%) | 177 ( 92.7%) |
| | Yes | 9 ( 8.9%) | 5 ( 5.6%) | 14 ( 7.3%) |
| DAY 9 | n | 100 | 89 | 189 |
| | No | 94 ( 94.0%) | 84 ( 94.4%) | 178 ( 94.2%) |
| | Yes | 6 ( 6.0%) | 5 ( 5.6%) | 11 ( 5.8%) |

Note:
Comparaisons at day 3,7,10 and 14 are considered for secondary analysis. The other days are used for exploratory analysis.


CONFIDENTIALITY STATEMENT: Information contained in this statistical report is confidential and should not be disclosed to other parties than those directly involved with the execution of this study without written authorization from Montreal Heart Institute

2 - Efficacy Analysis
2.2 - Secondary Analysis
Table 2.2.3k - Summary of and Statistical Analysis for Presence of Irritability/Confusion at Day 1 to 14, ITT population

| | | Placebo N=109 | Hesperidin N=107 | All N=216 |
|---|---|---|---|---|
| DAY 10 | n | 100 | 90 | 190 |
| | No | 96 ( 96.0%) | 86 ( 95.6%) | 182 ( 95.8%) |
| | Yes | 4 ( 4.0%) | 4 ( 4.4%) | 8 ( 4.2%) |
| DAY 11 | n | 98 | 86 | 184 |
| | No | 93 ( 94.9%) | 82 ( 95.3%) | 175 ( 95.1%) |
| | Yes | 5 ( 5.1%) | 4 ( 4.7%) | 9 ( 4.9%) |
| DAY 12 | n | 99 | 85 | 184 |
| | No | 97 ( 98.0%) | 82 ( 96.5%) | 179 ( 97.3%) |
| | Yes | 2 ( 2.0%) | 3 ( 3.5%) | 5 ( 2.7%) |
| DAY 13 | n | 98 | 83 | 181 |
| | No | 96 ( 98.0%) | 80 ( 96.4%) | 176 ( 97.2%) |
| | Yes | 2 ( 2.0%) | 3 ( 3.6%) | 5 ( 2.8%) |
| DAY 14 | n | 95 | 79 | 174 |
| | No | 94 ( 98.9%) | 76 ( 96.2%) | 170 ( 97.7%) |
| | Yes | 1 ( 1.1%) | 3 ( 3.8%) | 4 ( 2.3%) |

Note:
Comparaisons at day 3,7,10 and 14 are considered for secondary analysis. The other days are used for exploratory analysis.


CONFIDENTIALITY STATEMENT: Information contained in this statistical report is confidential and should not be disclosed to other parties than those directly involved with the execution of this study without written authorization from Montreal Heart Institute

2 – Efficacy Analysis
2.2 – Secondary Analysis
Table 2.2.3k - Summary of and Statistical Analysis for Presence of Irritability/Confusion at Day 1 to 14, ITT Population (continued)

Generalized linear mixed model (repeated binary logistic regression) with terms for treatment group, time and treatment group x time interaction

| Contrast | OR (95% CI) | P-value |
|---|---|---|
| Hesperidin vs Placebo at Day 3 | 1.15 (0.57; 2.34) | 0.6963 |
| Hesperidin vs Placebo at Day 7 | 0.65 (0.22; 1.88) | 0.4250 |
| Hesperidin vs Placebo at Day 10 | 1.12 (0.27; 4.65) | 0.8797 |
| Hesperidin vs Placebo at Day 14 | 3.71 (0.37; 37.03) | 0.2634 |


CONFIDENTIALITY STATEMENT: Information contained in this statistical report is confidential and should not be disclosed to other parties than those directly involved with the execution of this study without written authorization from Montreal Heart Institute

2 - Efficacy Analysis
2.2 - Secondary Analysis
Table 2.2.31 - Summary of and Statistical Analysis for Presence of Diarrhea at Day 1 to 14, ITT population

| | | Placebo N=109 | Hesperidin N=107 | All N=216 |
|---|---|---|---|---|
| DAY 1 | n | 107 | 105 | 212 |
| | No | 85 ( 79.4%) | 82 ( 78.1%) | 167 ( 78.8%) |
| | Yes | 22 ( 20.6%) | 23 ( 21.9%) | 45 ( 21.2%) |
| DAY 2 | n | 106 | 105 | 211 |
| | No | 84 ( 79.2%) | 83 ( 79.0%) | 167 ( 79.1%) |
| | Yes | 22 ( 20.8%) | 22 ( 21.0%) | 44 ( 20.9%) |
| DAY 3 | n | 103 | 102 | 205 |
| | No | 87 ( 84.5%) | 80 ( 78.4%) | 167 ( 81.5%) |
| | Yes | 16 ( 15.5%) | 22 ( 21.6%) | 38 ( 18.5%) |
| DAY 4 | n | 103 | 96 | 199 |
| | No | 90 ( 87.4%) | 72 ( 75.0%) | 162 ( 81.4%) |
| | Yes | 13 ( 12.6%) | 24 ( 25.0%) | 37 ( 18.6%) |
| DAY 5 | n | 102 | 95 | 197 |
| | No | 89 ( 87.3%) | 76 ( 80.0%) | 165 ( 83.8%) |
| | Yes | 13 ( 12.7%) | 19 ( 20.0%) | 32 ( 16.2%) |
| DAY 6 | n | 103 | 92 | 195 |
| | No | 92 ( 89.3%) | 72 ( 78.3%) | 164 ( 84.1%) |
| | Yes | 11 ( 10.7%) | 20 ( 21.7%) | 31 ( 15.9%) |
| DAY 7 | n | 102 | 91 | 193 |
| | No | 92 ( 90.2%) | 76 ( 83.5%) | 168 ( 87.0%) |
| | Yes | 10 ( 9.8%) | 15 ( 16.5%) | 25 ( 13.0%) |
| DAY 8 | n | 101 | 90 | 191 |
| | No | 92 ( 91.1%) | 77 ( 85.6%) | 169 ( 88.5%) |
| | Yes | 9 ( 8.9%) | 13 ( 14.4%) | 22 ( 11.5%) |
| DAY 9 | n | 100 | 89 | 189 |
| | No | 92 ( 92.0%) | 81 ( 91.0%) | 173 ( 91.5%) |
| | Yes | 8 ( 8.0%) | 8 ( 9.0%) | 16 ( 8.5%) |

Note:
Comparaisons at day 3,7,10 and 14 are considered for secondary analysis. The other days are used for exploratory analysis.


CONFIDENTIALITY STATEMENT: Information contained in this statistical report is confidential and should not be disclosed to other parties than those directly involved with the execution of this study without written authorization from Montreal Heart Institute

2 - Efficacy Analysis
2.2 - Secondary Analysis
Table 2.2.31 - Summary of and Statistical Analysis for Presence of Diarrhea at Day 1 to 14, ITT population

| | | Placebo N=109 | Hesperidin N=107 | All N=216 |
|---|---|---|---|---|
| DAY 10 | n | 100 | 90 | 190 |
| | No | 94 ( 94.0%) | 84 ( 93.3%) | 178 ( 93.7%) |
| | Yes | 6 ( 6.0%) | 6 ( 6.7%) | 12 ( 6.3%) |
| DAY 11 | n | 98 | 86 | 184 |
| | No | 94 ( 95.9%) | 79 ( 91.9%) | 173 ( 94.0%) |
| | Yes | 4 ( 4.1%) | 7 ( 8.1%) | 11 ( 6.0%) |
| DAY 12 | n | 99 | 85 | 184 |
| | No | 96 ( 97.0%) | 77 ( 90.6%) | 173 ( 94.0%) |
| | Yes | 3 ( 3.0%) | 8 ( 9.4%) | 11 ( 6.0%) |
| DAY 13 | n | 98 | 83 | 181 |
| | No | 94 ( 95.9%) | 76 ( 91.6%) | 170 ( 93.9%) |
| | Yes | 4 ( 4.1%) | 7 ( 8.4%) | 11 ( 6.1%) |
| DAY 14 | n | 95 | 79 | 174 |
| | No | 91 ( 95.8%) | 75 ( 94.9%) | 166 ( 95.4%) |
| | Yes | 4 ( 4.2%) | 4 ( 5.1%) | 8 ( 4.6%) |

Note:
Comparaisons at day 3,7,10 and 14 are considered for secondary analysis. The other days are used for exploratory analysis.


CONFIDENTIALITY STATEMENT: Information contained in this statistical report is confidential and should not be disclosed to other parties than those directly involved with the execution of this study without written authorization from Montreal Heart Institute

2 – Efficacy Analysis
2.2 – Secondary Analysis
Table 2.2.3l - Summary of and Statistical Analysis for Presence of Diarrhea at Day 1 to 14, ITT Population (continued)

Generalized linear mixed model (repeated binary logistic regression) with terms for treatment group, time and treatment group x time interaction

| Contrast | OR (95% CI) | P-value |
|---|---|---|
| Hesperidin vs Placebo at Day 3 | 1.50 (0.73; 3.06) | 0.2710 |
| Hesperidin vs Placebo at Day 7 | 1.82 (0.77; 4.30) | 0.1748 |
| Hesperidin vs Placebo at Day 10 | 1.12 (0.34; 3.63) | 0.8513 |
| Hesperidin vs Placebo at Day 14 | 1.21 (0.29; 5.07) | 0.7906 |


CONFIDENTIALITY STATEMENT: Information contained in this statistical report is confidential and should not be disclosed to other parties than those directly involved with the execution of this study without written authorization from Montreal Heart Institute

2 - Efficacy Analysis
2.2 - Secondary Analysis
Table 2.2.3m - Summary of and Statistical Analysis for Presence of Anosmia at Day 1 to 14, ITT Population

| | | Placebo N=109 | Hesperidin N=107 | All N=216 |
|---|---|---|---|---|
| DAY 1 | n | 107 | 105 | 212 |
| | No | 57 ( 53.3%) | 62 ( 59.0%) | 119 ( 56.1%) |
| | Yes | 50 ( 46.7%) | 43 ( 41.0%) | 93 ( 43.9%) |
| DAY 2 | n | 106 | 105 | 211 |
| | No | 52 ( 49.1%) | 56 ( 53.3%) | 108 ( 51.2%) |
| | Yes | 54 ( 50.9%) | 49 ( 46.7%) | 103 ( 48.8%) |
| DAY 3 | n | 103 | 102 | 205 |
| | No | 44 ( 42.7%) | 50 ( 49.0%) | 94 ( 45.9%) |
| | Yes | 59 ( 57.3%) | 52 ( 51.0%) | 111 ( 54.1%) |
| DAY 4 | n | 103 | 96 | 199 |
| | No | 44 ( 42.7%) | 48 ( 50.0%) | 92 ( 46.2%) |
| | Yes | 59 ( 57.3%) | 48 ( 50.0%) | 107 ( 53.8%) |
| DAY 5 | n | 102 | 95 | 197 |
| | No | 45 ( 44.1%) | 45 ( 47.4%) | 90 ( 45.7%) |
| | Yes | 57 ( 55.9%) | 50 ( 52.6%) | 107 ( 54.3%) |
| DAY 6 | n | 103 | 92 | 195 |
| | No | 52 ( 50.5%) | 43 ( 46.7%) | 95 ( 48.7%) |
| | Yes | 51 ( 49.5%) | 49 ( 53.3%) | 100 ( 51.3%) |
| DAY 7 | n | 102 | 91 | 193 |
| | No | 55 ( 53.9%) | 48 ( 52.7%) | 103 ( 53.4%) |
| | Yes | 47 ( 46.1%) | 43 ( 47.3%) | 90 ( 46.6%) |
| DAY 8 | n | 101 | 90 | 191 |
| | No | 60 ( 59.4%) | 50 ( 55.6%) | 110 ( 57.6%) |
| | Yes | 41 ( 40.6%) | 40 ( 44.4%) | 81 ( 42.4%) |
| DAY 9 | n | 100 | 89 | 189 |
| | No | 61 ( 61.0%) | 51 ( 57.3%) | 112 ( 59.3%) |
| | Yes | 39 ( 39.0%) | 38 ( 42.7%) | 77 ( 40.7%) |

Note:
Comparaisons at day 3,7,10 and 14 are considered for secondary analysis. The other days are used for exploratory analysis.


CONFIDENTIALITY STATEMENT: Information contained in this statistical report is confidential and should not be disclosed to other parties than those directly involved with the execution of this study without written authorization from Montreal Heart Institute

2 - Efficacy Analysis
2.2 - Secondary Analysis
Table 2.2.3m - Summary of and Statistical Analysis for Presence of Anosmia at Day 1 to 14, ITT Population

| | | Placebo N=109 | Hesperidin N=107 | All N=216 |
|---|---|---|---|---|
| DAY 10 | n | 100 | 90 | 190 |
| | No | 63 ( 63.0%) | 56 ( 62.2%) | 119 ( 62.6%) |
| | Yes | 37 ( 37.0%) | 34 ( 37.8%) | 71 ( 37.4%) |
| DAY 11 | n | 98 | 86 | 184 |
| | No | 63 ( 64.3%) | 55 ( 64.0%) | 118 ( 64.1%) |
| | Yes | 35 ( 35.7%) | 31 ( 36.0%) | 66 ( 35.9%) |
| DAY 12 | n | 99 | 85 | 184 |
| | No | 65 ( 65.7%) | 58 ( 68.2%) | 123 ( 66.8%) |
| | Yes | 34 ( 34.3%) | 27 ( 31.8%) | 61 ( 33.2%) |
| DAY 13 | n | 98 | 83 | 181 |
| | No | 65 ( 66.3%) | 58 ( 69.9%) | 123 ( 68.0%) |
| | Yes | 33 ( 33.7%) | 25 ( 30.1%) | 58 ( 32.0%) |
| DAY 14 | n | 95 | 79 | 174 |
| | No | 64 ( 67.4%) | 59 ( 74.7%) | 123 ( 70.7%) |
| | Yes | 31 ( 32.6%) | 20 ( 25.3%) | 51 ( 29.3%) |

Note:
Comparaisons at day 3,7,10 and 14 are considered for secondary analysis. The other days are used for exploratory analysis.


CONFIDENTIALITY STATEMENT: Information contained in this statistical report is confidential and should not be disclosed to other parties than those directly involved with the execution of this study without written authorization from Montreal Heart Institute

2 – Efficacy Analysis
2.2 – Secondary Analysis
Table 2.2.3m - Summary of and Statistical Analysis for Presence of Anosmia at Day 1 to 14, ITT Population (continued)

Generalized linear mixed model (repeated binary logistic regression) with terms for treatment group, time and treatment group x time interaction

| Contrast | OR (95% CI) | P-value |
|---|---|---|
| Hesperidin vs Placebo at Day 3 | 0.78 (0.45; 1.35) | 0.3686 |
| Hesperidin vs Placebo at Day 7 | 1.05 (0.59; 1.86) | 0.8711 |
| Hesperidin vs Placebo at Day 10 | 1.03 (0.57; 1.87) | 0.9124 |
| Hesperidin vs Placebo at Day 14 | 0.70 (0.36; 1.37) | 0.2952 |


CONFIDENTIALITY STATEMENT: Information contained in this statistical report is confidential and should not be disclosed to other parties than those directly involved with the execution of this study without written authorization from Montreal Heart Institute
