## Supplementary material for "Fourteen-days Evolution of COVID-19 Symptoms During the Third Wave in Non-vaccinated Subjects and Effects of Hesperidin Therapy: A randomized, double-blinded, placebo-controlled study": S4 Table

**S4 Table. Overall summary of treatment emergent adverse events (TEAE) and serious adverse events (TESAE), safety population.**

|  | **Placebo**  **N = 108** | **Hesperidin**  **N = 107** | **All**  **N = 215** |
| --- | --- | --- | --- |
| **Total number of TEAEs reported** | 16 | 23 | 39 |
| **Subjects with at least one TEAE** | 16 (14.8%) | 20 (18.7%) | 36 (16.7%) |
| **Subjects with at least one severe TEAE** | 2 (1.9%) | 3 (2.8%) | 5 (2.3%) |
| **Subjects with at least one TEAE related to the study treatment** | 4 (3.7%) | 3 (2.8%) | 7 (3.3%) |
| **Subjects with at least one TEAE leading to drug withdrawal** | 5 (4.6%) | 8 (7.5%) | 13 (6.0%) |
| **Total number of TESAEs reported** | 1 | 5 | 6 |
| **Subjects with at least one TESAE** | 1 (0.9%) | 4 (3.7%) | 5 (2.3%) |
| **Subjects with at least one severe TESAE** | 1 (0.9%) | 3 (2.8%) | 4 (1.9%) |
| **Subjects with at least one TESAE related to the study treatment** | 0 (0%) | 0 (0%) | 0 (0%) |
| **Subjects with at least one TEAE leading to drug withdrawal** | 1 (0.9%) | 2 (1.9%) | 3 (1.4%) |
